# Transmission of SARS-CoV-2 associated with aircraft travel: a systematic review (Version 1)

**DOI:** 10.1101/2021.06.03.21258274

**Authors:** EC Rosca, C Heneghan, EA Spencer, J Brassey, A Plüddemann, IJ Onakpoya, D Evans, JM Conly, T Jefferson

## Abstract

**Background:** Air travel may be associated with the spread of viruses via infected passengers and potentially through in-flight transmission. Given the novelty of the SARS-CoV-2 virus, transmission associated with air travel is based on what is known about the dynamics of transmission of other respiratory virus infections, especially those due to other coronaviruses and influenza. Our objective was to provide a rapid summary and evaluation of relevant data on the transmission of SARS-CoV-2 aboard aircraft, report important policy implications, and highlight research gaps requiring urgent attention.

**Methods:** This review is part of an Open Evidence Review on Transmission Dynamics of SARS-CoV-2. We searched LitCovid, medRxiv, Google Scholar, and the WHO Covid-19 database from 1 February 2020 to 27 January 2021 and included studies on the transmission of SARS-CoV-2 aboard aircraft. We assessed study quality based on five criteria and reported important findings.

**Results:** We included 18 studies on in-flight transmission of SARS-CoV-2, representing 130 unique flights and two studies on wastewater from aircraft. The overall quality of reporting was low. Two wastewater studies reported PCR-positive SARS-CoV-2 samples, but with relatively high Cycle threshold values ranging from 36 to 40. The definition of an index case was very heterogeneous across the studies. The proportion of contacts traced ranged from 0.68% to 100%. In total, the authors successfully traced 2800/19729 passengers, 140/180 crew members, and 8/8 medical staff. Altogether, 273 index cases were reported, with 64 secondary cases. No secondary cases were reported in three studies, each investigating one flight. The secondary attack rate among the studies that followed up >80% of the passengers and crew (including data on 10 flights) varied between 0% and 8.2%. The included studies reported on the possibility of SARS-CoV-2 transmission from asymptomatic, pre-symptomatic, and symptomatic individuals. Viral cultures were performed in two studies, with 10 positive results reported. Genomic sequencing and phylogenetic analysis were performed in individuals from four flights, with the completeness of genomic similarity ranging from 81-100%.

**Conclusion:** Current evidence suggests that SARS-CoV-2 can be transmitted during aircraft travel, but the published data do not permit any conclusive assessment of the likelihood and extent. Furthermore, the quality of evidence from most published studies is low. The variation in study design and methodology restricts the comparison of findings across studies. Standardized guidelines for conducting and reporting future studies of transmission on aircrafts should be developed.

## Introduction

SARS-CoV-2 is a new coronavirus strain that spreads rapidly. The World Health Organization (WHO), national governments, and public health officials have been working to coordinate the response and rapid development of prevention, control, and management measures on several fronts. The overarching aim is to control COVID-19 by suppressing transmission of the virus and prevent associated illness and death. [WHO 2020] However, the transmission of the SARS-CoV-2 virus and the many facets of the illness it causes are incompletely understood, and public health measures for restricting transmission are based on best available information [EASA 2020].

Air travel may be associated with the spread of viruses via infected passengers and potentially through in-flight transmission. The high number of passengers, frequently in close proximity to each other, increases the likelihood of transmitting infectious diseases via microorganisms which may be spread through multiple routes of transmission. As in other closed/semi-closed settings, the on-board transmission of viruses can be facilitated by direct person-to-person contact, contact with contaminated surfaces [ECDC 2014; Leitmeyer 2011; Leitmeyer & Adlhoch 2016] and droplet transmission. The risk of transmission of infections depends on contact among passengers at the departure gate, proximity to an index case, passengers, crew movement, and fomites [ECDC 2017; Hertzberg & Weiss 2016].

The WHO and the European Centre for Disease Prevention and Control (ECDC) have elaborated specific guidance recommendations for case management in air transport for several pathogens [ECDC, 2014; WHO, 2009].

Given the novelty of SARS-CoV-2, air travel transmission models of spread are based on what is known of the dynamics of other respiratory infections, especially those due to other coronaviruses and influenza. One of the most important aspects of models of spread is the uncertainty regarding the modes and circumstances of transmission of newly identified agents. Consequently, research is ongoing to understand SARS-CoV-2 modes of transmission, with a continuous array of new publications. As a result, there is a need to continuously and systematically conduct reviews of available studies with the latest knowledge to inform recommendations using the most up-to-date information.

## Objectives

Our objectives were to provide a rapid summary and evaluation of relevant data on the transmission of SARS-CoV-2 aboard aircraft, report important policy implications, and highlight research areas urgently needed. This transmission area includes airborne, contact and droplet, fomite, and orofecal.

## Methods

The present work is an open evidence review on the transmission of SARS-CoV-2 in aircraft. The protocol (Appendix 1) was developed based on a previous protocol for a series of systematic reviews on the evidence on transmission dynamics of COVID-19 (Appendix 2) (Jefferson et al. 2020) (see https://www.cebm.net/evidence-synthesis/transmission-dynamics-of-covid-19/ for the original protocol).

For this review we conducted searches in the following electronic databases: LitCovid, medRxiv, Google Scholar, and the WHO Covid-19 database up to 27 January 2021. Search terms were *Covid-19, SARS-CoV-2, transmission*, and *airplane* appropriate synonyms (Appendix 3). In addition, we screened for additional studies the reference lists of relevant articles, including reviews and the systematic review on close contact transmission of SARS-CoV-2 [Onakpoya 2021]. We did not impose any language restrictions.

We included studies reporting on the on-board transmission of SARS-CoV-2, from passengers and crew to passengers or crew. We considered any potential transmission mode, including droplet, airborne, fomite, fecal-oral, or other. We included studies of any design, except predictive or modelling studies.

From the included studies, we extracted the following information: publication details (authors, year, country); study type; flight characteristics (origin and destination of the aircraft, flight duration, technical specifications of the airplane, ground delays, and information on ventilation systems); data on the index cases (number, age, gender, country of residence or nationality, seating, whether they wore masks, symptoms during the flight, laboratory confirmation of diagnosis); details on contact tracing (definition of contact, secondary cases demographic data, symptoms, laboratory confirmation, contact tracing strategy, methods used to identify contacts, methods used for contacting contacts, total number of contacts identified, the total number of successfully traced contacts, the seating of contacts in relation to the index case, immunological status and if they wore or not masks); exposure of primary and secondary cases (before, during, and after the flight); conclusion on disease transmission (the number of cases/number of contacted passengers, and crew excluding index cases), interventions used, and source of funding for the study. One reviewer (ECR) extracted data from the included studies, and these were independently checked by a second reviewer (CH).

We assessed the quality of included human studies on a modified QUADAS-2 tool using five criteria: (1) a clearly defined setting (aircraft details, location of index cases and secondary cases), (2) demographic characteristics (age, gender), sampling procedures adequately described with the day of the sampling procedure and data on symptoms (with onset day); (3) follow-up duration sufficient for the outcomes; (4) the transmission outcomes assessed adequately (including demographic, clinical and paraclinical data); (5) main biases that are threats to validity taken into consideration (follow up > 80% of individuals, alternative exposures excluded) (Appendix 1). For non-human studies, we used the modified QUADAS-2 tool to assess the following aspects: (1) description of methods with sufficient detail to replicate, (2) sample sources clear, (3) analysis and reporting appropriate, (4) bias assessment, and (5) applicability (Appendix1).

The QUADAS-2 tool was adapted because the included studies were not primarily designed as diagnostic accuracy studies. One reviewer (ECR) assessed the reporting quality of included studies and these were independently checked by a second reviewer (EAS). Disagreements were resolved by consensus.

For studies that generated hypothesis testing of on-board COVID-19 transmission, we also assessed the strength of evidence of each study depending on the methods used to investigate the SARS-CoV-2 transmission (Jefferson 2021). We presented the results in tabular format. We reported results of specific subgroups of studies where relevant. The included studies showed substantial heterogeneity; therefore, we considered meta-analyses inappropriate.

## Results

Our searches identified 753 studies out of which 20 were considered eligible (see Figure 1). We assessed in fulltext 25 studies. We excluded five studies: two narrative reviews, two modeling studies, and one preprint version of an included study (Appendix 4). In total, we included 20 studies: two studies on the wastewater from aircrafts [Ahmed 2020; Albastaki 2021]; and 18 studies considering in-flight transmission of SARS-CoV-2 [Bae 2020, Chen 2020; Choi 2020; Eldin 2020; Hoehl 2020; Khanh 2020; Kong 2020; Murphy 2020; Ng 2020; Nir-Paz 2020; Park 2020; Pavli 2020; Schwartz 2020; Speake 2020; Swadi 2021; Yang 2020; J. Zhang 2020; X. A. Zhang 2020] (Appendix 5).

**Figure 1.**
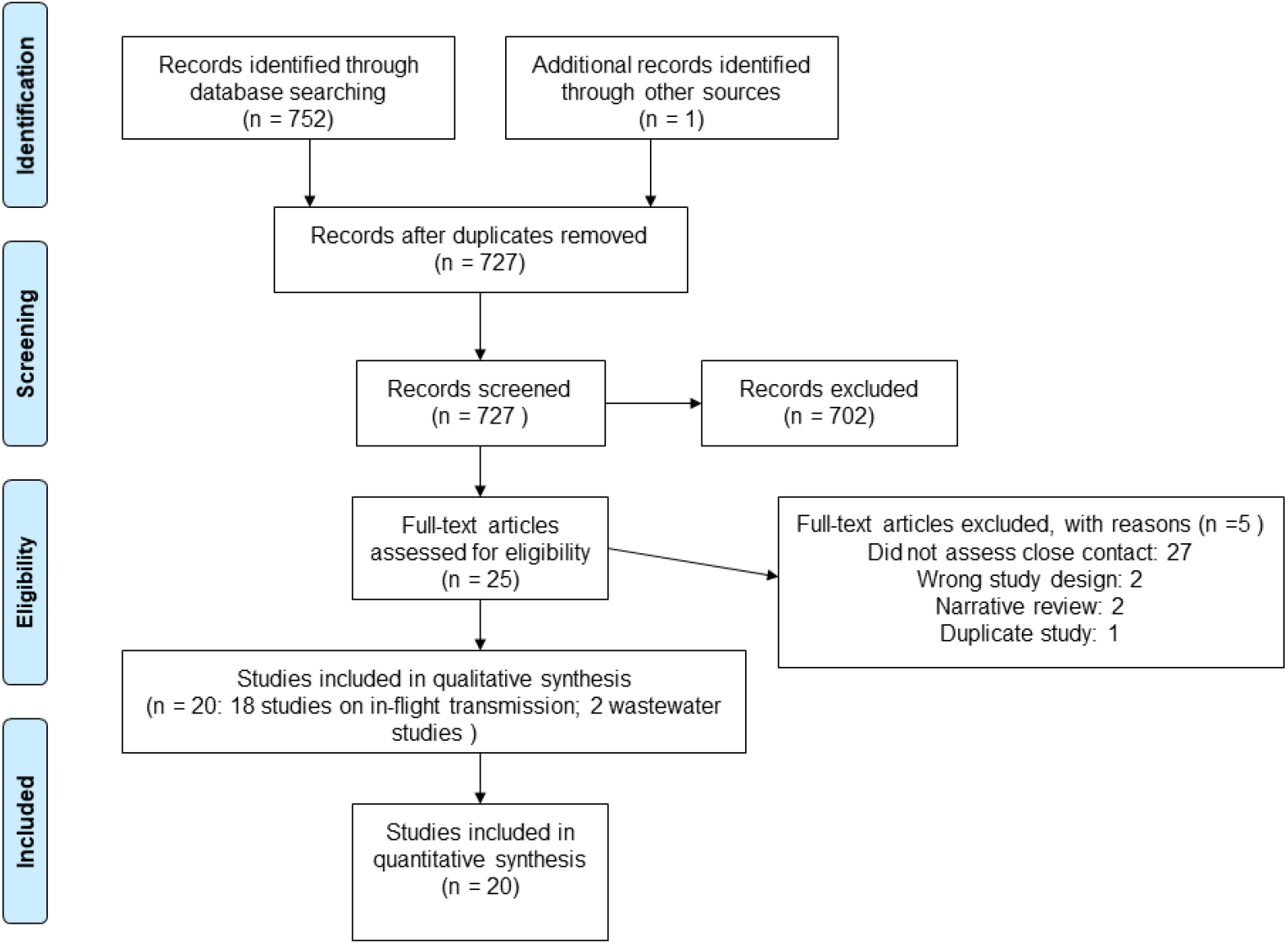
Flow diagram showing the process for inclusion of studies assessing aircraft transmission of SARS-CoV-2.

The main characteristics of the included studies are presented in Table 1 and Table 2.

**Table 1.** In-flight transmission studies.

| Study | Year | Country | No of passengers and crew | No of index cases | No of passengers and crew traced (%) | No of secondary cases identified (%) | Attack rate (%) | No of secondary cases within 2 rows (%) | No of secondary cases outside the area of 2 rows (%) | Strength of evidence |
| --- | --- | --- | --- | --- | --- | --- | --- | --- | --- | --- |
| Bae | 2020 | South Korea | 299 px; 10 crew; 8 medical staff | 6 px | 299 px; 10 crew; 8 medical staff (100%) | 1 px | 1/311 (0.32%) | 0 | 1 | RT-PCR, no data on Ct |
| Chen | 2020 | China | 335 px; 11 crew | 15 px | 335 px; 11 crew (100%) | 1 px | 1/331 (0.30%) | 1 | 0 | RT-PCR, no data on Ct |
| Choi | 2020 | China | 294 px; unknown no of crew members | 2 px | 2 crew (only symptomatic COVID-19 cases and known contacts of those and other cases were identified and traced) (1.36%) | 2 crew | N/A | N/A | N/A | RT-PCR, no data on Ct, **GS** |
| Eldin | 2020 | France | Not specified | 1 px | 3 px | 1 px | N/A | Not specified | Not specified | RT-PCR, no data on Ct |
| Hoehl | 2020 | Germany | 102 px | 7 px | 71 px (69.60%) | 2 px | 2/71 | 2 px | 0 | RT-PCR, no data on Ct; SARS-CoV-2 IgG |
| Khanh | 2020 | Vietnam | 201 px; 16 crew | 1 px | 168 px; 16 crew (83.40%) | 14px; 1 crew | 15/184 (8.15%) | 11 px | 1 px; 1 crew | RT-PCR, no data on Ct |
| Kong - flight 1 | 2021 | China | 59 px | 1 px | 58 px (98.3%) | 3 px (unclear) | N/A | 0 | 3 px | RT-PCR, no data on Ct |
| Kong - flight 2 | 2021 | China | 232 px | Unspecified | 232 px (100%) | 2 px | N/A | Not specified | Not specified | Symptomatic, exposure to COVID-19; RT-PCR negative, no data on Ct |
| Murphy | 2020 | Ireland | 49 px; 12 crew | Unknown (1 to 7) | 37 px (60.65%) | 4 to 12 | 4/41 (9.8%) to 12/48 (25%) | Not specified | Not specified | **GS 5/13 px** |
| Ng | 2020 | Singapore | 94 px | 2 px | 92 px (96.66%) | 1 (1.08%) | 1/92 (1.08%) | Not specified | Not specified | RT-PCR, no data on Ct |
| Nir-Paz | 2020 | Israel | 11 px | 2 px | 11 px | 0 | 0 | 0 | 0 | index cases - RT-PCR positive (Ct 34 and Ct 24), positive **viral cultures.** |
| Park | 2020 | Korea | Not specified | 30 px | Not specified | 1 crew | N/A | N/A | N/A | RT-PCR positive, Ct <40 |
| Pavly - flight 1 | 2020 | Greece | 164px; 6 crew | 2 px | 163 px, 6 crew (99.41%) | 4 px, 1 crew | 5/167 (2.99%) | 4 px, 1 crew | 0 | RT-PCR, no data on Ct |
| Pavly b - flights 2 | 2020 | Greece | 2203px, 110 crew | 21 px | 870px, 90 crew (41.50%) | 4 px, 1 crew | 5/960 | 4 px, 1 crew | 0 | RT-PCR, no data on Ct |
| Schwartz | 2020 | Canada | approx. 350 px, unspecified number of crew | 1 | Unclear | 0 | N/A | 0 | 0 | RT-PCR, no data on Ct |
| Speake | 2020 | Australia | 241 px | 18 px | 46 px (19.8%) | 8 px | 8/223 | 5 px | 3 px | RT-PCR; no data on Ct. **GS; Viral cultures** |
| Swadi | 2021 | New Zealand | 86 px | 2 px | 86 px (100%) | 4 px | 4/84 (4.76%) | 4 | 0 | RT-PCR, with Ct data; **GS** |
| Yang | 2020 | China | 325 px | 1 px | 9 px, 9 crew (5.53%) | 9 px | N/A | Not specified | Not specified | RT-PCR, no data on Ct |
| Zhang XA - flight 1 | 2020 | China | Unspecified | 2 px | 8 px, 1 crew | 0 px | N/A | 0 | 0 | Epidemiologic data; RT-PCR negative in 1 px; RT-PCR not done in asymptomatic cases (5px, 1 crew); No data on Ct |
| Zhang XA - flight 2 | 2020 | China | 343 px, 21 crew | Unclear; minimum 3 px | 325 px, 11 crew (92.3%) | Unclear, max 7 px | N/A | Not specified | Not specified | RT-PCR, No data on Ct |
| Zhang J | 2020 | China | 14505 | 159 px | 161 px (1.10%) | 2 px | 0.14 ‰ | Not specified | Not specified | RT-PCR, no data on Ct |

*Abbreviations: px – passengers; Ct - cycle threshold; RT-PCR - real time reverse transcription–polymerase chain reaction; GS – genome sequencing.*

**Table 2.**
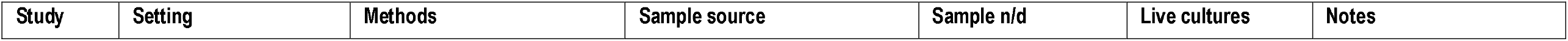
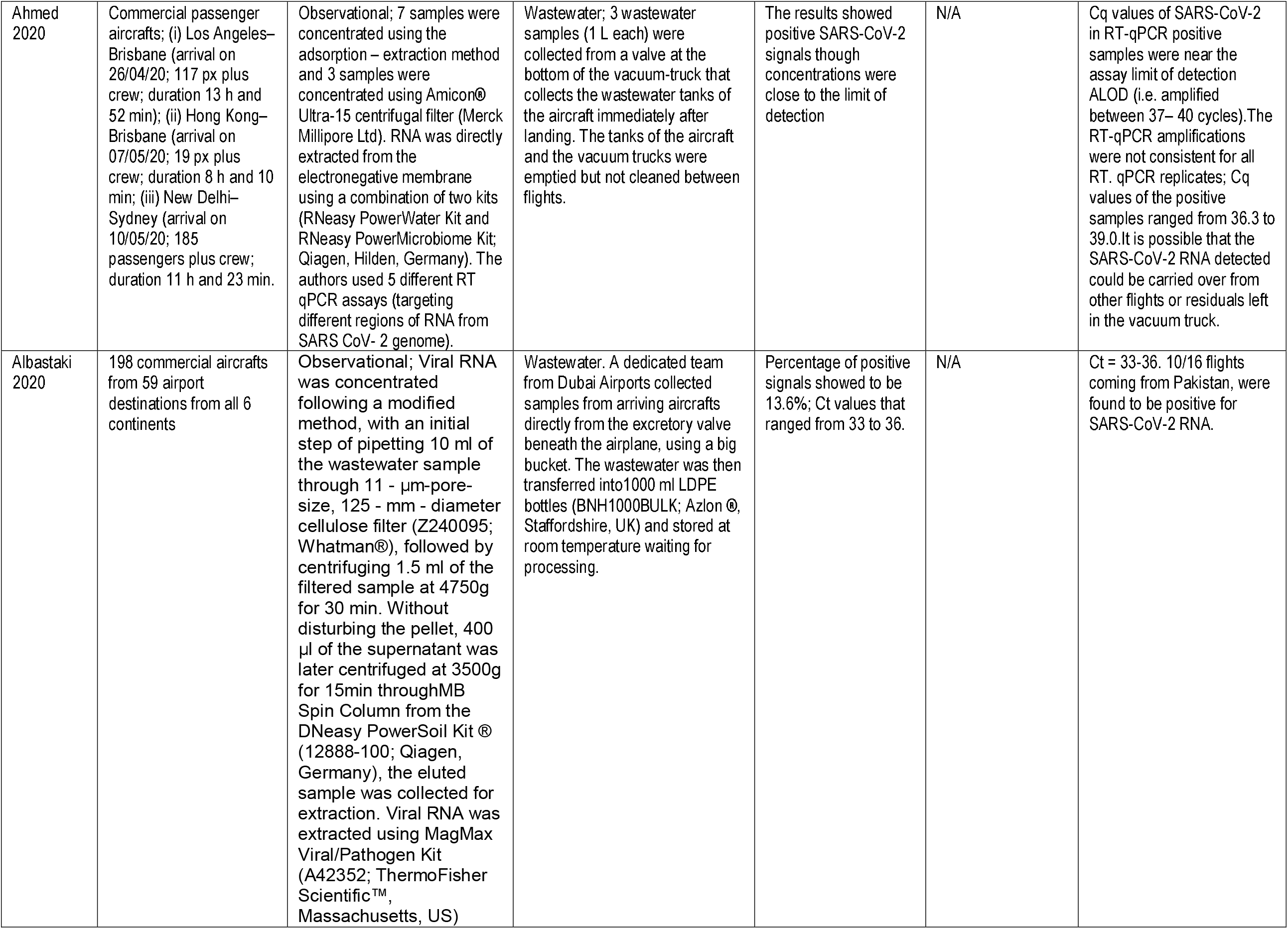
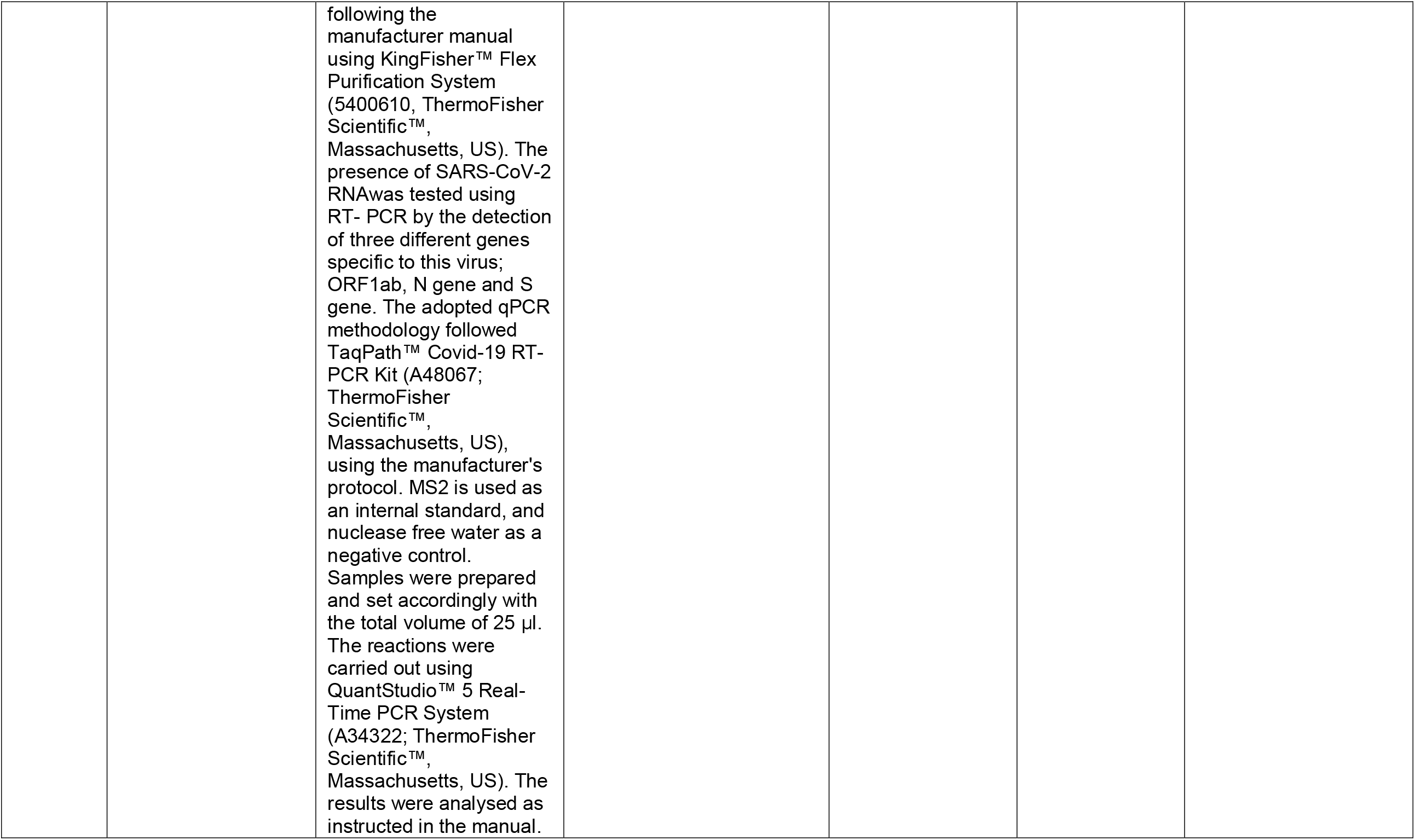
Non-human studies (wastewater studies).

In the present review, we included two studies that were very likely based on the same flight [Chen 2020; Yang 2020]. Despite our efforts to clarify this issue (e.g., contacting the authors and the editors of the journals), we could not ascertain with certainty whether both studies report the same flight. Although there are many similarities between the results of the studies, there are also some minor discrepancies, including the number of passengers (335 vs. 325), the number of index cases (15 vs.1) and the arrival time (9:40 pm vs. 10:00 pm). We considered it to be highly unusual to have 2 flights arriving within 20 minutes of each other with full passenger loads with the same departure and arrival sites. What is most important is the one investigation reporting 15 cases and suggesting in-flight transmission and the other suggesting that the cases were incubating SARS-CoV-2 from community acquisition and, if the same flight, illustrates how dramatically different conclusions were reached between the two investigations. A detailed comparison of the data extracted from the studies is presented in Appendix 6.

### Quality of included studies

None of the included studies reported a published protocol. The risk of bias assessment of the included studies is presented in Table 3a and Table 3b.

**Table 3.a.** Quality assessment of included studies: in-flight transmission studies.

| Study | Study type | Clearly defined setting | Demographic characteristics / sampling procedures adequately described | Follow-up strategy and duration sufficient for the outcomes | The transmission outcomes assessed adequately | Main threats to validity taken into consideration? | Notes |
| --- | --- | --- | --- | --- | --- | --- | --- |
| Bae 2020 | Cohort, prospective | Yes | Unclear | Yes | Unclear | Yes | No data on Ct |
| Chen 2020 | Cohort, prospective | Yes | Unclear | Yes | Unclear | Unclear | No data on Ct; case ascertainment, alternative exposures, recall bias |
| Choi 20220 | Retrospective, case series | Yes | Unclear | No | Unclear | No | No data on Ct; alternative exposures; asymptomatic px may be missed; use of GISAID |
| Eldin 2020 | Retrospective, case study | No | Unclear | No | Unclear | No | No data on Ct; no comprehensive contact tracing (<20%); use of a database (which may be incomplete); alternative exposures |
| Hoehl 2020 | Cohort, retrospective | Yes | Unclear | No | Unclear | No | No data on Ct; use of other laboratory measures (IgG); tested only symptomatic px; > 20% of px were not tested |
| Khanh 2020 | Cohort, retrospective | Yes | Unclear | Yes | Unclear | Yes | No data on Ct; |
| Kong 2021- flight 1 | Cohort, retrospective | Yes | Unclear | No | Unclear | No | No data on Ct; asymptomatic cases may be missed; recall bias; alternative exposures; unclear number of secondary cases; |
| Kong 2021 - flight 2 | Cohort, retrospective | Yes | Unclear | No | Unclear | No | No data on Ct; asymptomatic cases may be missed; recall bias; alternative exposures; |
| Murphy 2020 | Cohort, retrospective | Yes | No | No | Unclear | No | Unclear number/data of index cases and secondary cases; no data on Ct; tracing –60.65%; alternative exposures; WGS in 5/13 |
| Ng 2020 | Cohort | No | Unclear | Yes | Unclear | No | No data on Ct; alternative exposure |
| Nir-Paz 2020 | Cohort | Yes | No | Yes | Unclear | Unclear | Ct – 34 (1 px, with no viral vultures); no data on crew |
| Park 2020 | Cohort, retrospective | No | No | No | Unclear | No | Index cases: upper respiratory tract (n = 23) Ct = 27.0 (22.1–32.0); Lower respiratory tract (n = 27) Ct = 26.4 (22.7–28.8); Secondary case: Ct< 40. no comprehensive tracing strategy; alternative exposures; |
| Pavly 2020 - flight 1 | Cohort, retrospective | No | Unclear | No | Unclear | No | No data on Ct; tracing strategy – within 2 seats; recall bias, alternative exposures |
| Pavly 2020 - all flights | Cohort, retrospective | No | Unclear | No | Unclear | No | No data on Ct; tracing strategy – within 2 seats, recall bias, |
| Schwartz 2020 | Cohort, retrospective | No | Unclear | No | Unclear | No | No data on Ct; asymptomatic cases could be missed; alternative exposures; no date of symptoms onset or RT-PCR |
| Speake 2020 | Cohort | Yes | Unclear | No | Unclear | No | No data on Ct; alternative exposures; asymptomatic cases could be missed; recall bias; use of GISAID |
| Swadi 2021 | Cohort, retrospective | Yes | Yes | Yes | Yes | No | Use of GISAID; some Ct > 25; alternative exposures |
| Yang 2020 | Case series, retrospective | No | Unclear | No | Unclear | No | No data on Ct; follow-up <20%; alternative exposure, case ascertainment |
| Zhang XA 2020 - flight 1 | Case series, retrospective | No | Unclear | No | Unclear | No | No data on Ct; follow-up – within 2 rows; asymptomatic cases were not tested; alternative exposure |
| Zhang XA 2020 - flight 2 | Cohort, retrospective | Yes | Unclear | No | Unclear | No | No data on Ct; follow-up -asymptomatic cases were not tested; alternative exposure |
| Zhang J 2020 | Cohort, retrospective | No | Unclear | No | Unclear | No | No data on Ct; follow-up < 20%; no data on RT-PCR date; no data on the date of the symptom's onset, asymptomatic cases were not tested, selection bias |

Abbreviations: Ct - cycle threshold; RT-PCR - real time reverse transcription–polymerase chain reaction.

**Table 3.b.** Quality assessment of included studies: in-flight transmission studies.

| Study | Study type | Description of methods with sufficient detail to replicate | Sample sources clear | Analysis and reporting appropriate | Is bias dealt with | Applicability | Notes |
| --- | --- | --- | --- | --- | --- | --- | --- |
| Ahmed 2020 | Observational | Yes | Yes | Yes | No | Yes | Cq values of the positive samples ranged from 36.3 to 39.0. |
| Albastaki 2020 | Observational | Yes | Yes | Yes | No | Yes | Ct of the positive samples = 33-36 |

For the two studies on wastewater [Ahmed 2020; Albastaki 2021] the description of methods with sufficient detail to replicate the findings was considered adequate. The sample sources were clear, the analysis and reporting were considered appropriate, and there were no concerns about their applicability. However, we considered no studies adequately addressed the potential biases (Table 3b).

Regarding the in-flight transmission studies, 12/130 flights (9.2%) presented a clearly defined setting, and 1/130 flights (0.77%) adequately described demographic characteristics and sampling procedures. In 6/130 (4.6%) flights, the strategy and duration of follow-up were found sufficient for the outcome assessments. The transmission outcomes were considered to be adequately assessed for only 1/130 (0.77%) flights, and data validity concerns were taken into consideration for 2/130 (1.54%) flights (Table 3a). The overall quality of the latter category of studies was considered low (Figure 2).

**Figure 2.**
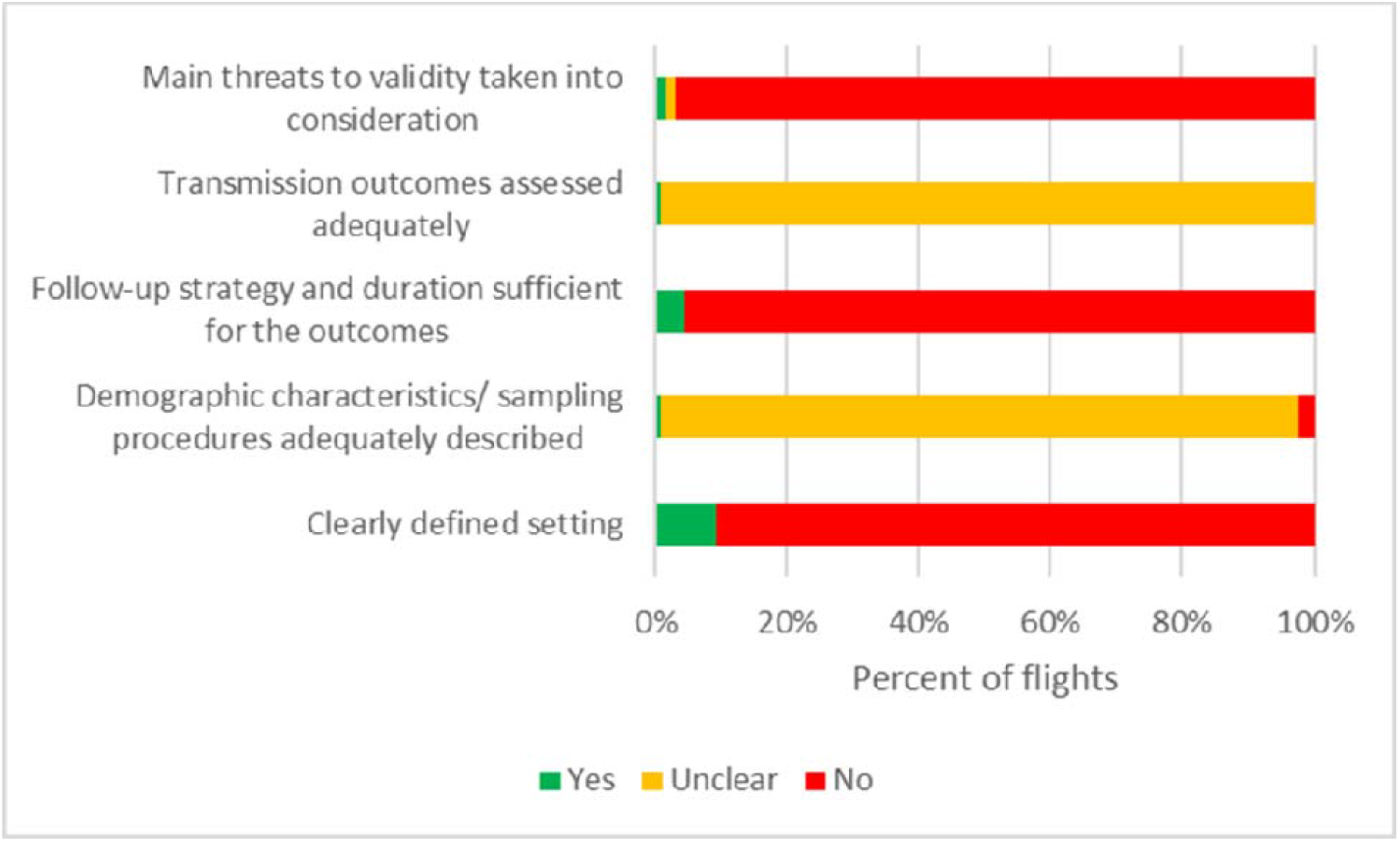
Risk of bias graph in studies on in-flight transmission of SARS-CoV-2.

### Wastewater studies

One study investigated the wastewater from three commercial passenger aircrafts [Ahmed 2020]. The results showed positive SARS-CoV-2 RNA in the samples processed though concentrations were close to the limit of detection (Cq values ranged from 36 to 39) (Table 2). The second study [Albastaki 2021] investigated the wastewater of 198 commercial aircrafts from 59 airport destinations from all 6 continents. The percentage of positive SARS-CoV-2 RNA was 13.6%, with Ct values ranging from 33 to 36 (Table 2).

### Studies on the in-flight transmission of SARS-CoV-2

#### Flight details

The total number of flights (n = 130) exceeded the number of the included studies (n = 18). Four studies reported multiple flights (See Appendix 7): two flights [Kong 2020; Zhang XA], eighteen flights [Pavli 2020], and 94 flights [Zhang J]. The aircraft type was reported for 11 flights [Chen 2020; Choi 2020; Hoehl 2020; Nir-Paz 2020; Speake 2020; Swadi 2021; J Zhang 2020], and the flight numbers were provided for six flights [Khanh 2020; Kong 2020; Park 2020; XA Zhang 2020]. No technical specifications data on the airplanes was mentioned for 113 flights. The flight duration was reported in 15 studies, ranging from approximately 2 to 18 hours. Nine flights were long duration, lasting more than 7 hours; one flight was short-haul, lasting about 2 hours, and 5 flights had a medium duration, between 3 to 5 hours. Flight time was not specified in 115 flights. Ground delays were not reported by any study except for one where the aircraft had a refueling stop of 2 hours, and the auxiliary power unit was inoperative for approximately 30 minutes, with inoperative ventilation [Swadi 2021]. Data on the ventilation system was provided for only three flights. Two studies reported on the airflow in the cabin [Hoehl 2020; J. Zhang 2020], and one study described the ventilation system [Nir-Paz 2020].

### Case definitions: index cases, contacts, and secondary cases

The definition of index case varied across the studies (see Appendix 7) and included asymptomatic, pre-symptomatic, and symptomatic individuals. A clear definition of the index cases was not provided for 103 flights, and the contact definitions differed between studies. Two studies, including 95 flights, did not provide any information on contacts. The case definitions for secondary infections were also variable, including asymptomatic and symptomatic passengers or crew.

### Study types and contact tracing strategies

The majority of included studies presented retrospective follow-up of passengers and crew after identifying one or more index cases (Table 1, Appendix 7). Some authors also used travel and airline information data, medical records from hospitals, telephone interviews, or a notifiable disease database. A prospective study with the immediate quarantine of all the passengers was done for 2 flights [Bae 2020; Ng 2020]. Active daily contact monitoring was done in one study [Schwartz 2020], and active surveillance with quarantine of > 80% of the passengers and crew was reported for one flight [Zhang XA 2020]. The time span for initiating follow-up ranged between the day of arrival and several months. The follow-up strategies focused on passengers seated within two rows or two meters of the index case, passengers in the same section or class, or used a comprehensive approach. In addition, crew members of 25 flights were followed up for possible transmission of SARS-CoV-2. The proportion of contacts identified and traced ranged from 0.68% [Choi 2020] to 100% [Bae 2020; Chen 2020].

In total, the authors identified 19,729 passengers, 180 crew members, and 8 medical staff. Among them, they successfully traced 2.800 passengers, 140 crew members, and 8 medical staff. Three studies did not report the number of passengers or crew members on the aircraft board [Eldin 2020; Park 2020; X. A. Zhang 2020].

### On-board transmission

Overall, 273 index cases were reported across 18 studies. However, three studies did not clearly report the number of index cases, and therefore, we considered the minimum number of index cases in each report [Eldin 2020; Murphy 2020; X. A. Zhang 2020].

In the index cases, laboratory diagnosis was based on RT-PCR in all 18 studies (Appendix 7). The RT-PCR timing varied from the day of arrival to day 11 after the flight, and passengers were pre-symptomatic, symptomatic, or asymptomatic. Fifteen studies report a binary result (positive/negative) for passengers or crew from 127 flights, including 239 cases [Bae 2020; Chen 2020; Choi 2020; Eldin 2020; Hoehl 2020; Khanh 2020; Kong 2020; Murphy 2020; Ng 2020; Pavli 2020; Schwartz 2020; Speake 2020; Yang 2020; J. Zhang 2020; X. A. Zhang 2020]. The RT-PCR Ct was reported for 34 index cases from three flights [Nir-Paz 2020; Park 2020; Swadi 2021].

Only three index cases reported in two studies had a positive RT-PCR test at a Ct value < 25 [Nir-Paz 2020; Swadi 2021]. One study reported that among the 30 index cases, 23 upper respiratory samples were positive at a median Ct of 27 (interquartile range 22.1-32.0), and 27 lower respiratory tract samples were positive at a median Ct of 26.4 (interquartile range 22.7 – 28.8) [Park 2020].

In total, 64 secondary cases were reported (59 passengers and 5 crew members). The number of secondary cases was not clear in 3 reports [Kong 2020; Murphy 2020; X. A. Zhang 2020]. Three studies, each investigating one flight, reported no secondary cases [Nir-Paz 2020; Schwartz 2020; Yang 2020]. The secondary attack rate (number of secondary cases / all successfully traced persons) among the studies that followed-up > 80% of the passengers and crew [Bae 2020; Chen 2020; Khanh 2020; Kong 2020; Ng 2020; Nir-Paz 2020; Pavli 2020; Swadi 2021; X.A. Zhang 2020], varied between 0% [Nir-Paz 2020] and 8.2% [Khanh 2020]. However, GS was performed only in one study that reported a secondary attack rate of 4.76% [Swadi 2020].

The secondary cases were asymptomatic or symptomatic individuals. The symptoms onset day ranged from the first day after arrival to the 24^th^ day after arrival. They presented an RT-PCR test positive for SARS-CoV-2. The sample collection timing ranged from the second to the 16^th^ day after arrival. In one study [Hoehl 2020], the diagnosis of secondary infection was based on SARS-CoV-2 IgG antibodies, performed on week 7 or week 9 after the flight.

The seating position of secondary cases in relation to the index cases was specified for 24 flights across eight studies [Bae 2020; Chen 2020; Hoehl 2020; Khanh 2020; Kong 2020; Pavli 2020; Speake 2020; Swadi 2021], with 27 passengers seated within 2 rows or 2 meters [Chen 2020; Hoehl 2020; Khanh 2020; Pavli 2020; Speake 2020; Swadi 2021] and one crew member who served the index cases [Pavly 2020], and four studies reported eight passengers seated outside of 2 rows (or presumed 2 meters) from the index cases, and one crew [Bae 2020; Khanh 2020; Kong 2020; Speake 2020). The seating position of the secondary cases was not specified in eight studies for 102 flights [Choi 2020; Eldin 2020; Murphy 2020; Ng 2020; Park 2020; Yang 2020; J. Zhang 2020; X. A. Zhang 2020].

Regarding the use of masks, one study reported the use of N95 masks [Bae 2020], one study reported the use of FFP2 masks [Nir-Paz 2020], while seven studies did not report on masking of passengers or crew [Choi 2020; Eldin 2020; Khanh 2020; Kong 2020; Park 2020; Pavli 2020; X. A. Zhang 2020].

Alternative exposures were not fully assessed in 13 studies including for 32 flights [Chen, 2020; Choi 2020; Eldin 2020; Kong 2020; Murphy 2020; Ng 2020; Park 2020; Pavli 2020; Schwartz 2020; Speake 2020; Swadi 2021; Yang 2020; X. A. Zhang 2020]. Furthermore, in three studies including 21 flights, some secondary cases were family members [Ng 2020; Pavli 2020; X. A. Zhang 2020].

In eleven studies, asymptomatic passengers or crew members from 106 flights were not tested for SARS-CoV-2 infection [Choi 2020; Eldin 2020; Hoehl 2020; Kong 2020; Ng 2020; Park 2020; Schwartz 2020; Speake 2020; Yang 2020; J. Zhang 2020; X. A. Zhang 2020).

### Genome sequencing (GS) and phylogenetic analysis

Genome sequencing and phylogenic analysis were performed in individuals from four flights [Choi 2020; Murphy 2020; Speake 2020; Swadi 2021]. The methods used for performing these investigations were essentially similar across the studies (see Table 4). The completeness of coverage of the positive samples ranged from 81-100% across the studies. The phylogenetic analysis showed more than 99% similarity across the entire viral genomes.

**Table 4.** Genomic sequencing studies.

| Study | Methods for genome sequencing | Phylogenetic analysis | Results |
| --- | --- | --- | --- |
| Choi 2020 | Authors analyzed 4 RT-PCR-confirmed COVID-19 cases on a flight (2 index cases, 2 secondary cases). Stored samples of these cases were then sent to a WHO reference laboratory at the University of Hong Kong for full genome analyses. Near full-length genomes (N≥ 29760 nucleotides) were deduced by Illumina sequencing method using the primers and protocol previously described by authors. All the deduced sequences had a minimum coverage of 100 or above. The specimens were sequenced and analyzed blind to the passenger/crew/case status of the four individuals. | Representative sequences from each phylogenetic clade of SARS-CoV-2 (G, GH, GR, L, O, S and V) were retrieved from GISAID. Viral sequences were aligned and phylogenetically analyzed using BioEdit and MEGA-X, respectively. A phylogenetic tree was constructed by the neighbor-joining method with bootstrap testing (N=1,000). Metadata from 191 Hong Kong viral sequences deposited in GISAID were also used in the analyses. | Authors investigated the near full-length genomes from two index cases and two secondary cases. The study revealed that they were 100% identical and phylogenetically grouped to the same clade; all deduced sequences had a minimum coverage of 100. |
| Murphy 2020 | GS in 5 of the 13 flight-associated cases of SARS-CoV-2 infection which came from one case travelling from one continent, three cases travelling from a different continent and one case travelling from a third continent. | All 5 samples were identified as belonging to SARS-CoV-2 viral lineage B.1.36 (PANGOLIN nomenclature, v2.0.7 | Pairwise comparison of the nucleotide sequences showed more than 99% homology across the entire viral genome, strongly suggesting a single point source of infection. |
| Speake 2020 | Processed reads were mapped to the SARS-CoV-2 reference genome (GenBank accession no. MN908947). Primer-clipped alignment files were imported into Geneious Prime version 2020.1.1 for coverage analysis before consensus calling, and consensus sequences were generated by using iVar version 1.2.2. | Genome sequences of SARS-CoV-2 from Western Australia were assigned to lineages by using the Phylogenetic Assignment of Named Global Outbreak Lineages (PANGOLIN) tool (https://github.com/covlineages/pangolin External Link). On July 17, 2020, authors retrieved SARS-CoV-2 complete genomes with corresponding metadata from the GISAID database. The final dataset contained 540 GISAID whole-genome sequences that were aligned with the sequences from Western Australia generated in this study by using MAFFT version 7.467. Phylogenetic trees were visualized in iTOL (Interactive Tree of Life, https://itol.embl.de External Link) and MEGA version 7.014. | 100% coverage was obtained for 21 and partial coverage (81%–99%) for 4 samples. The phylogenetic tree for the 21 complete genomes belonged to either the A.2 (n = 17) or B.1 (n = 4) sub-lineages of SARS-CoV-2 |
| Swadi 2021 | Independent viral extracts were prepared by the Institute of Environmental Science and Research (Porirua, New Zealand) from the 7 positive respiratory tract samples in which SARS-CoV-2 was initially detected by RRT-PCR. Authors extracted RNA from SARS-CoV-2-positive samples and subjected it to whole-genome sequencing by following the 1,200-bp amplicon protocol and Oxford Nanopore Rapid barcoding R9.0 sequencing. Genomic data are available on GISAID | The lineage of the genomes obtained from the 7 passengers was determined by using pangolin version 2.0.8 (https://pangolin.cog-uk.io) and compared with genomes from the same lineage available on GISAID. Genomes were aligned by using MAFFT version 7 (8) and using the FFT-NS-2 progressive alignment algorithm. Authors estimated a maximum likelihood phylogenetic tree by using IQ-TREE version 1.6.8 and the Hasegawa-Kishino-Yano nucleotide substitution model, with a gamma distributed rate variation among sites (HKY+Γ), the best-fit model as determined by ModelFinder, and branch support assessment by using the ultrafast bootstrap method. | All SARS-CoV-2 samples from the 7 passengers were subjected to whole-genome sequencing for surveillance purposes. The sequences obtained were assigned to lineage B.1 and were genetically identical, apart from 1 mutation for the sample from passenger D. By comparing these 7 genomes to the international database (GISAID), authors identified 6 additional identical genomes: 4 from Switzerland and 2 from the United Kingdom, sampled during September 2–23. These findings were consistent with virus introduction onto the airplane from Switzerland by passenger A, B, or both. Nevertheless, accurately identifying the source of this outbreak may be impeded by substantial biases and gaps in global sequencing data. Hence, authors cannot explicitly exclude passenger C as the source. |

One study investigating the near full-length genomes from two index cases and two secondary cases found that they were 100% identical and phylogenetically grouped to the same clade; all deduced sequences had a minimum coverage of 100 [Choi 2020].

In another study, the authors performed GS in 5 of the 13 flight-associated cases of SARS-CoV-2 infection. They found 99% homology across the entire virus genome in all 5 cases [Murphy 2020].

A third study [Speake 2020] reported that sufficient viral RNA was available to generate an adequate sequence for 25 of the 29 samples that were RT-PCR positive. The authors obtained 100% coverage for 21 and partial coverage (81%–99%) for 4 samples. The phylogenetic tree for the 21 complete genomes revealed that they belonged to either the A.2 (n = 17) or B.1 (n = 4) sublineages of SARS-CoV-2. All of the complete A.2 sequences belonged to a distinct genomic cluster separated by <2 single-nucleotide polymorphisms. The 4 B.1 viruses comprised 3 B.1.31 and 1 phylogenetically more distant B.1 strain. Of the 4 partial sequences, 3 clustered with the A2 strains, and the other was designated B.1.1 and was phylogenetically close to the B.1.31 sequences [Speake 2020].

On another flight, the authors demonstrated that the viral sequences of the index cases and the secondary cases were assigned to lineage B.1 and were genetically identical, apart from 1 mutation from the sample from one secondary case [Swadi 202]). Three studies used databases (e.g., GISAID) to identify the country of the source of infection [Choi 2020; Speake 2020; Swadi 2021].

### Viral cultures

Two studies [Nir-Paz 2020; Speake 2020] performed viral culture (Table 1, Table 5, Appendix 7). One study [Nir-Paz., 2020] reported that one asymptomatic index case presented positive viral cultures 4 days after arrival. No data was provided on the methods used for the cultures. The Ct value of RT-PCR, performed on the first day after arrival was 24. The authors report that the passenger presented an RT-PCR positive for 26 days, but the latter Ct values are not specified [Nir-Paz 2020].

**Table 5.** Viral cultures.

| Study | Participants | Methods | Results |
| --- | --- | --- | --- |
| Nir-Paz 2020 | 1 asymptomatic index case | Not reported | Viral cultures positive 4 days after arrival |
| Speake 2020 | 17 RT-PCR-positive specimens | Virus culture was attempted for primary samples. Clinical specimens were inoculated in triplicate wells with Vero-E6 cells at 80% confluency, incubated at 37°C in 5% CO<sub>2</sub>, and inspected for cytopathic effect daily for up to 10 days. Identity was confirmed by in-house PCRs. | The authors attempted to culture 17 PCR-positive specimens, nine (53%) of which grew SARS-CoV-2. Of note, 4/11 persons who were infectious on the flight had culture-positive specimens collected the next day. 11 passengers had PCR-confirmed SARS-CoV-2 infection and symptom onset within 48 hours of the flight. They had been in the same cabin with symptomatic persons who had culture-positive A2-RP virus strain. |

In another study [Speake 2020], viral cultures were performed using Vero E6 cells. Specimens were inspected for cytopathic effects daily for up to 10 days. The authors attempted to culture 17 PCR-positive specimens, nine (53%) of which grew SARS-CoV-2. Of note, 4/11 persons who were infectious on the flight had culture-positive specimens collected the next day.

## Discussion

### Summary of main findings

We identified 18 studies assessing the in-flight transmission of SARS-CoV-2 and 2 studies investigating the presence of the virus in the wastewater of aircrafts. The evidence from the studies reporting on the on-board transmission suggests that the risk of infection could be higher in individuals seated within 2 rows of the index cases. Nonetheless, identifying secondary cases seated within a greater distance limits the evidence for restricting the contact tracing to this area.

Regarding the duration of the flight, there were short, medium, and long flights with a low or a high number of secondary cases. For example, in one short flight, of approximately two hours, the authors reported two index cases and five secondary cases [Pavli 2020]. Another study investigating a flight that lasted 18 hours reported two index cases and four secondary cases [Swadi 2021]. The hypothesis on the assumption that the risk of transmission increases with the length of flight due to higher exposure postulated in other airborne diseases [Dowdall 2010] needs further investigation.

It is not clear whether the use of masks can prevent transmission of SARS-CoV-2 in flights. The flights where some of the passengers and crew used FFP2 masks [Nir-Paz 2020] or N95 masks [Bae 2020] presented an attack rate of 0% and 0.32% respectively. However, the authors did not specify if a “fit test” was performed to assess if the mask fits and seals properly so potentially contaminated air cannot leak into the respirator. Furthermore, most of the studies did not provide clear data on the masking of passengers and crew.

The included studies reported on the possibility of SARS-CoV-2 transmission from asymptomatic, pre-symptomatic, or symptomatic individuals. However, a major limitation of most studies consisted of the possibility of asymptomatic index cases transmitting the infection and of asymptomatic secondary cases not being investigated due to lack of any symptoms, with lowering the quality of case ascertainment.

In addition, the number of studies that reported on Ct of RT-PCR is limited; therefore, case ascertainments are likely to be biased [Jefferson 2021]. The timeline of the sample collections also is suggestive of bias in some studies.

The four studies that performed GS and phylogenetic analysis [Choi 2020; Murphy 2020; Speake 2020; Swadi 2021] report higher quality reliable evidence, indicating that aircraft may be a setting associated with SARS-CoV-2 transmission. GS alone cannot prove the presence of infectious materials, as the amplicon-based methods now often used to assemble SARS-CoV-2 genome sequences require only viral RNA. Nor can amplification-based SARS-CoV-2 sequencing exclude infections caused by other agents such as rhinoviruses and OC43. However, such methods do provide secure phylogenetic insights into the relationship between the putative index and secondary cases. Nonetheless, the use of databases like Global initiative on sharing all influenza data (GISAID) to ascertain transmission may induce bias. A recent review [Furuse, 2021] found that, even though many developing countries have high numbers of SARS-CoV-2 infected cases, they have few published sequences. Such missing data could create bias in a phylogeographic analysis to elucidate the global transmission dynamics of SARS-CoV-2. Substantial gaps in global sequencing data may impede the accurate identification of a source of infection.

The positive results of viral cultures observed in two studies [Nir-Paz 2020; Speake 2020] bring further evidence on aircraft transmission of SARS-CoV-2. The positive viral cultures of index cases indicate that infectious virus was present, with potential for transmission to the secondary cases. The transmission of the virus to the secondary cases is documented by the evidence that the index case was contaminated (i.e., Ct values <25) with infectious virus (i.e., cultivatible e virus); the spread is confirmed by genetic sequencing, associated with the proof that they were clearly exposed to the virus in the environment (i.e., the route of transmission). It is noteworthy that samples from the environment were not performed in any of the studies.

Nonetheless, the authors of one study [Nir-Paz 2020] did not report on the methods used for viral cultures, and they did not perform a GS. They report non-transmission of SARS-CoV-2 to the other passengers of the flight, based on RT-PCR results, but the Ct values of the other individuals are not provided.

The second study [Speake 2020] had an elegant design with WGS and viral cultures. They did not provide Cts which would have provided additional insights into the relative abundance of infectious materials in the environment.

Similar to previous studies on aircraft transmission of pathogens [Leitmeyer & Adlhoch 2016], the validity of many studies is limited by the possibility of alternative exposures. Some common sites of alternative exposures include sites before the flight (i.e., waiting spaces), during flight (i.e., at the lavatory, movement of passengers during flight), and after landing (i.e., lining up to exit the aircraft, security checkpoints, documents check).

The variations observed in the contact tracing strategies, the timeliness of contact tracing, the proportion of passengers and crew successfully traced, the use of different case definitions, the testing strategy, and case ascertainment also give rise to further doubts about the validity of the overall findings.

Our review results are consistent with the suggestion that transmission of SARS-CoV-2 can occur in aircrafts but is a relatively rare event. Similar to the close contact transmission [Onakpoya 2021a], the research shows evidence of positive virus cultures as well as genomic evidence of on-board transmission in one study [Speake 2020]. However, the route of transmission needs further investigation. For example, recent systematic reviews reported a lack of positive viral cultures in studies on airborne [Heneghan 2021 a] and fomite transmission [Onakpoya 2021b]. Up to date, positive viral cultures were demonstrated only by studies on orofecal transmission [Heneghan 2021b] transmission.

This review did not address a comparison of risk between aircraft and non-aircraft settings. Furthermore, there is currently little evidence on the risk for transmission within comparable, non-aircraft settings (i.e. enclosed spaces like theaters or subways), with air exchange filtration system, mask wearing and minimal movement once in place, with variable screening strategies before entry.

To our knowledge, no other systematic review of the literature has been undertaken to assess the evidence for transmission of SARS-CoV-2 aboard aircraft. We performed an extensive search of the literature for eligible studies, accounted for the quality of included studies, and have reported outcomes (GS and viral cultures). We included results from one non-peer-reviewed study, which may affect the reliability of the review results. However, due to the ongoing pandemic, such studies could potentially be of research benefit.

The limitations in this review are mainly related to the quality of the included studies and the fact that we could not ascertain with certainty if two papers were reporting on the same flight [Chen 2020; Yang 2020]. In addition, the data extraction was challenging due to missing, incomplete, or unclear descriptions of the investigations. In addition, we may not have identified all relevant studies examining the SARS-CoV-2 transmission of aircraft associated transmission events.

Our findings emphasize the need for a standardized approach to investigation and reporting on the transmission of SARS-CoV-2 aboard aircraft.

Future studies should aim for comprehensive assessment of passengers and crew, with a complete follow-up strategy. Factors that may influence transmission, such as infectivity of the index case (asymptomatic, pre-symptomatic, or symptomatic, with or without mask), the susceptibility of passengers (previous COVID-19 infection or vaccination, wearing or not of masks), and effectiveness of exposure (proximity to the index case, duration of exposure, technical specifications of the airplane, quality of cabin air) should be consistently assessed across studies.

Research should include Ct values when reporting RT-PCR results and describe the timing and sample collection methods. In addition, further studies, including virus isolation, whole-genome sequencing, and phylogenetic analysis, should be conducted to strengthen the current evidence. Therefore, standardization of research reporting should be a priority. Furthermore, new studies should take into account other factors that might impact transmission patterns, including natural immunity and vaccination coverage.

### Conclusion

Current evidence indicates that the risk of transmission of SARS-CoV-2 aboard aircraft is low, but the published data do not permit any conclusive assessment of the likelihood and extent. Furthermore, the quality of evidence from most published studies is low. The variation in study design and methodology restricts the analysis of findings across studies. Standardized guidelines for the reporting of future research should be developed.

## Data Availability

All data included in the review are provided in the tables or in the supplemental files.

## Competing Interests

TJ was in receipt of a Cochrane Methods Innovations Fund grant to develop guidance on the use of regulatory data in Cochrane reviews (2015-018). In 2014–2016, he was a member of three advisory boards for Boehringer Ingelheim. TJ was a member of an independent data monitoring committee for a Sanofi Pasteur clinical trial on an influenza vaccine. TJ is occasionally interviewed by market research companies about phase I or II pharmaceutical products for which he receives fees (current). TJ was a member of three advisory boards for Boehringer Ingelheim (2014-16). TJ was a member of an independent data monitoring committee for a Sanofi Pasteur clinical trial on an influenza vaccine (2015-2017). TJ is a relator in a False Claims Act lawsuit on behalf of the United States that involves sales of Tamiflu for pandemic stockpiling. If resolved in the United States favor, he would be entitled to a percentage of the recovery. TJ is coholder of a Laura and John Arnold Foundation grant for development of a RIAT support centre (2017-2020) and Jean Monnet Network Grant, 2017-2020 for The Jean Monnet Health Law and Policy Network. TJ is an unpaid collaborator to the project Beyond Transparency in Pharmaceutical Research and Regulation led by Dalhousie University and funded by the Canadian Institutes of Health Research (2018-2022). TJ consulted for Illumina LLC on next generation gene sequencing (2019-2020). TJ was the consultant scientific coordinator for the HTA Medical Technology programme of the Agenzia per i Servizi Sanitari Nazionali (AGENAS) of the Italian MoH (2007-2019). TJ is Director Medical Affairs for BC Solutions, a market access company for medical devices in Europe. TJ was funded by NIHR UK and the World Health Organization (WHO) to update Cochrane review A122, Physical Interventions to interrupt the spread of respiratory viruses. TJ is funded by Oxford University to carry out a living review on the transmission epidemiology of COVID-19. Since 2020, TJ receives fees for articles published by The Spectator and other media outlets. TJ is part of a review group carrying out Living rapid literature review on the modes of transmission of SARS-CoV-2 (WHO Registration 2020/1077093-0). He is a member of the WHO COVID-19 Infection Prevention and Control Research Working Group for which he receives no funds. TJ is funded to co-author rapid reviews on the impact of Covid restrictions by the Collateral Global Organisation. He is also an editor of the Cochrane Acute Respiratory Infections Group.

TJ’s competing interests are also online https://restoringtrials.org/competing-interests-tom-jefferson

CH holds grant funding from the NIHR, the NIHR School of Primary Care Research, the NIHR BRC Oxford and the World Health Organization for a series of Living rapid review on the modes of transmission of SARs-CoV-2 reference WHO registration No2020/1077093. He has received financial remuneration from an asbestos case and given legal advice on mesh and hormone pregnancy tests cases. He has received expenses and fees for his media work including occasional payments from BBC Radio 4 Inside Health and The Spectator. He receives expenses for teaching EBM and is also paid for his GP work in NHS out of hours (contract Oxford Health NHS Foundation Trust). He has also received income from the publication of a series of toolkit books and for appraising treatment recommendations in non-NHS settings. He is Director of CEBM, an NIHR Senior Investigator and an advisor to Collateral Global. He is also an editor of the Cochrane Acute Respiratory Infections Group.

DHE holds grant funding from the Canadian Institutes for Health Research and Li Ka Shing Institute of Virology relating to the development of Covid-19 vaccines as well as the Canadian Natural Science and Engineering Research Council concerning Covid-19 aerosol transmission. He is a recipient of World Health Organization and Province of Alberta funding which supports the provision of BSL3-based SARS-CoV-2 culture services to regional investigators. He also holds public and private sector contract funding relating to the development of poxvirus-based Covid-19 vaccines, SARS-CoV-2-inactivation technologies, and serum neutralization testing.

JMC holds grants from the Canadian Institutes for Health Research on acute and primary care preparedness for COVID 19 in Alberta, Canada and was the primary local Investigator for a Staphylococcus aureus vaccine study funded by Pfizer for which all funding was provided only to the University of Calgary. He is a co investigator on a WHO funded study using integrated human factors and ethnography approaches to identify and scale innovative IPC guidance implementation supports in primary care with a focus on low resource settings and using drone aerial systems to deliver medical supplies and PPE to remote First Nations communities during the COVID 19 pandemic. He also received support from the Centers for Disease Control and Prevention (CDC) to attend an Infection Control Think Tank Meeting. He is a member of the WHO Infection Prevention and Control Research and Development Expert Group for COVID 19 and the WHO Health Emergencies Programme (WHE) Ad hoc COVID 19 IPC Guidance Development Group, both of which provide multidisciplinary advice to the WHO, for which no funding is received and from which no funding recommendations are made for any WHO contracts or grants. He is also a member of the Cochrane Acute Respiratory Infections Group.

JB is a major shareholder in the Trip Database search engine (www.tripdatabase.com) as well as being an employee. In relation to this work Trip has worked with a large number of organisations over the years, none have any links with this work. The main current projects are with AXA and Collateral Global.

AP is Senior Research Fellow at the Centre for Evidence-Based Medicine and reports grant funding from NIHR School of Primary Care Research (NIHR SPCR ESWG project 390 and project 461), during the conduct of the study; and occasionally receives expenses for teaching Evidence-Based Medicine.

ECR was a member of the European Federation of Neurological Societies (EFNS) / European Academy of Neurology (EAN) Scientist Panel – Subcommittee of Infectious Diseases (2013-2017). Since 2021, she is a member of the International Parkinson and Movement Disorder Society (MDS) Multiple System Atrophy Study Group and the Mild Cognitive Impairment in Parkinson Disease Study Group. She was an External Expert and sometimes Rapporteur for COST proposals (2013, 2016, 2017, 2018, 2019) for Neurology projects.

IJO, and EAS have no interests to disclose.

## Data Availability

All data included in the review are provided in the tables or in the supplemental files.

## References

Ahmed, W., Bertsch, P. M., Angel, N., Bibby, K., Bivins, A., Dierens, L., Edson, J., Ehret, J., Gyawali, P., Hamilton, K. A., Hosegood, I., Hugenholtz, P., Jiang, G., Kitajima, M., Sichani, H. T., Shi, J., Shimko, K. M., Simpson, S. L., Smith, W. J. M., Symonds, E. M., Thomas, K. V., Verhagen, R., Zaugg, J., & Mueller, J. F. (2020). Detection of SARS-CoV-2 RNA in commercial passenger aircraft and cruise ship wastewater: a surveillance tool for assessing the presence of COVID-19 infected travelers. Journal of Travel Medicine, 27(5). https://doi.org/10.1093/jtm/taaa116

Albastaki, A., Naji, M., Lootah, R., Almeheiri, R., Almulla, H., Almarri, I., Alreyami, A., Aden, A., & Alghafri, R. (2021). First confirmed detection of SARS-COV-2 in untreated municipal and aircraft wastewater in Dubai, UAE: The use of wastewater based epidemiology as an early warning tool to monitor the prevalence of COVID-19. Sci Total Environ, 760, 143350. https://doi.org/10.1016/j.scitotenv.2020.143350

Bae, S. H., Shin, H., Koo, H. Y., Lee, S. W., Yang, J. M., & Yon, D. K. (2020). Asymptomatic Transmission of SARS-CoV-2 on Evacuation Flight. Emerg Infect Dis, 26(11), 2705–2708. https://doi.org/10.3201/eid2611.203353

Chen, J., He, H., Cheng, W., Liu, Y., Sun, Z., Chai, C., Kong, Q., Sun, W., Zhang, J., Guo, S., Shi, X., Wang, J., Chen, E., & Chen, Z. (2020). Potential transmission of SARS-CoV-2 on a flight from Singapore to Hangzhou, China: An epidemiological investigation. Travel Medicine and Infectious Disease, 36, 101816. https://doi.org/10.1016/j.tmaid.2020.101816

Choi, E. M., Chu, D. K. W., Cheng, P. K. C., Tsang, D. N. C., Peiris, M., Bausch, D. G., Poon, L. L. M., & Watson-Jones, D. (2020). In-Flight Transmission of SARS-CoV-2. Emerg Infect Dis, 26(11), 2713–2716. https://doi.org/10.3201/eid2611.203254

Dowdall, N. P., Evans, A. D., & Thibeault, C. (2010). Air Travel and TB: an airline perspective. Travel Med Infect Dis, 8(2), 96–103. https://doi.org/10.1016/j.tmaid.2010.02.006

EASA. (2020). COVID-19 Aviation Health Safety Protocol. Operational guidelines for the management of air passengers and aviation personnel in relation to the COVID-19 pandemic. Retrieved 3/30/2021 from EASA-ECDC_COVID-19_Operational guidelines for management of passengers_v2.pdf (europa.eu)

ECDC. (2014). Risk assessment guidelines for infectious diseases transmitted on aircraft (RAGIDA) - Influenza. European Centre for Disease Prevention and Control. Retrieved 3/23 from https://www.ecdc.europa.eu/en/publications-data/risk-assessment-guidelines-infectious-diseases-transmitted-aircraft-ragida#no-link

ECDC (2017). European Centre for Disease Prevention and Control. Infectious diseases on aircraft. Retrieved 3/30/2021 from https://www.ecdc.europa.eu/en/all-topics-z/travellers-health/infectious-diseases-aircraft

Eldin, C., Lagier, J.-C., Mailhe, M., & Gautret, P. (2020). Probable aircraft transmission of Covid-19 in-flight from the Central African Republic to France. Travel Medicine and Infectious Disease, 35, 101643. https://doi.org/10.1016/j.tmaid.2020.101643

Furuse, Y. (2021). Genomic sequencing effort for SARS-CoV-2 by country during the pandemic. Int J Infect Dis, 103, 305–307. https://doi.org/10.1016/j.ijid.2020.12.034

Heneghan CJ, Spencer EA, Brassey J et al. SARS-CoV-2 and the role of airborne transmission: a systematic review [version 1; peer review: approved with reservations, 2 not approved]. F1000Research 2021, 10:232 (https://doi.org/10.12688/f1000research.52091.1)

Heneghan CJ, Spencer EA, Brassey J et al. SARS-CoV-2 and the role of orofecal transmission: a systematic review [version 1; peer review: 1 approved with reservations]. F1000Research 2021, 10:231 (https://doi.org/10.12688/f1000research.51592.1)

Hertzberg, V.S., Weiss, H. (2016). On the 2-Row Rule for Infectious Disease Transmission on Aircraft. Ann Glob Health. 2016, 82(5):819–823. https://doi.org/10.1016/j.aogh.2016.06.003.

Hoehl, S., Karaca, O., Kohmer, N., Westhaus, S., Graf, J., Goetsch, U., & Ciesek, S. (2020). Assessment of SARS-CoV-2 Transmission on an International Flight and Among a Tourist Group. JAMA Netw Open, 3(8), e2018044. https://doi.org/10.1001/jamanetworkopen.2020.18044

Jefferson, T., Heneghan, C., Spencer, E., Brassey, J., Pluddeman, A., Onakpoya, I., Evans, D., Conly, J. A Hierarchical Framework for Assessing Transmission Causality of Respiratory Viruses. Preprints 2021, 2021040633 (doi: 10.20944/preprints202104.0633.v1).

Khanh, N. C., Thai, P. Q., Quach, H. L., Thi, N. H., Dinh, P. C., Duong, T. N., Mai, L. T. Q., Nghia, N. D., Tu, T. A., Quang, N., Quang, T. D., Nguyen, T. T., Vogt, F., & Anh, D. D. (2020). Transmission of SARS-CoV 2 During Long-Haul Flight. Emerg Infect Dis, 26(11), 2617–2624. https://doi.org/10.3201/eid2611.203299

Kong, D., Wang, Y., Lu, L., Wu, H., Ye, C., Wagner, A. L., Yang, J., Zheng, Y., Gong, X., Zhu, Y., Jin, B., Xiao, W., Mao, S., Jiang, C., Lin, S., Han, R., Yu, X., Cui, P., Fang, Q., Lu, Y., & Pan, H. (2020). Clusters of 2019 coronavirus disease (COVID-19) cases in Chinese tour groups. Transbound Emerg Dis. https://doi.org/10.1111/tbed.13729

Leitmeyer, K. (2011). European risk assessment guidance for infectious diseases transmitted on aircraft--the RAGIDA project. Euro Surveill, 16(16).

Leitmeyer, K., & Adlhoch, C. (2016). Review Article: Influenza Transmission on Aircraft: A Systematic Literature Review. Epidemiology, 27(5), 743–751. https://doi.org/10.1097/ede.0000000000000438

Murphy, N., Boland, M., Bambury, N., Fitzgerald, M., Comerford, L., Dever, N., O’Sullivan, M. B., Petty-Saphon, N., Kiernan, R., Jensen, M., & O’Connor, L. (2020). A large national outbreak of COVID-19 linked to air travel, Ireland, summer 2020. Euro Surveill, 25(42). https://doi.org/10.2807/1560-7917.Es.2020.25.42.2001624

Ng, O. T., Marimuthu, K., Chia, P. Y., Koh, V., Chiew, C. J., De Wang, L., Young, B. E., Chan, M., Vasoo, S., Ling, L. M., Lye, D. C., Kam, K. Q., Thoon, K. C., Kurupatham, L., Said, Z., Goh, E., Low, C., Lim, S. K., Raj, P., Oh, O., Koh, V. T. J., Poh, C., Mak, T. M., Cui, L., Cook, A. R., Lin, R. T. P., Leo, Y. S., & Lee, V. J. M. (2020). SARS-CoV-2 Infection among Travelers Returning from Wuhan, China. N Engl J Med, 382(15), 1476–1478. https://doi.org/10.1056/NEJMc2003100

Nir-Paz, R., Grotto, I., Strolov, I., Salmon, A., Mandelboim, M., Mendelson, E., & Regev-Yochay, G. (2020). Absence of in-flight transmission of SARS-CoV-2 likely due to use of face masks on board. Journal of Travel Medicine, 27(8). https://doi.org/10.1093/jtm/taaa117

Onakpoya IJ, Heneghan CJ, Spencer EA et al. SARS-CoV-2 and the role of close contact in transmission: a systematic review [version 1; peer review: awaiting peer review]. F1000Research 2021, 10:280 (https://doi.org/10.12688/f1000research.52439.1)

Onakpoya IJ, Heneghan CJ, Spencer EA et al. SARS-CoV-2 and the role of fomite transmission: a systematic review [version 1; peer review: 1 approved with reservations]. F1000Research 2021, 10:233 (https://doi.org/10.12688/f1000research.51590.1)

Park, J. H., Jang, J. H., Lee, K., Yoo, S. J., & Shin, H. (2020). COVID-19 Outbreak and Presymptomatic Transmission in Pilgrim Travelers Who Returned to Korea from Israel. J Korean Med Sci, 35(48), e424. https://doi.org/10.3346/jkms.2020.35.e424

Pavli, A., Smeti, P., Hadjianastasiou, S., Theodoridou, K., Spilioti, A., Papadima, K., Andreopoulou, A., Gkolfinopoulou, K., Sapounas, S., Spanakis, N., Tsakris, A., & Maltezou, H. C. (2020). In-flight transmission of COVID-19 on flights to Greece: An epidemiological analysis. Travel Med Infect Dis, 38, 101882. https://doi.org/10.1016/j.tmaid.2020.101882

Schwartz, K. L., Murti, M., Finkelstein, M., Leis, J. A., Fitzgerald-Husek, A., Bourns, L., Meghani, H., Saunders, A., Allen, V., & Yaffe, B. (2020). Lack of COVID-19 transmission on an international flight. Canadian Medical Association journal, 192(15), E410. https://doi.org/10.1503/cmaj.75015

Speake, H., Phillips, A., Chong, T., Sikazwe, C., Levy, A., Lang, J., Scalley, B., Speers, D. J., Smith, D. W., Effler, P., & McEvoy, S. P. (2020). Flight-Associated Transmission of Severe Acute Respiratory Syndrome Coronavirus 2 Corroborated by Whole-Genome Sequencing. Emerg Infect Dis, 26(12), 2872–2880. https://doi.org/10.3201/eid2612.203910

Swadi, T., Geoghegan, J. L., Devine, T., McElnay, C., Sherwood, J., Shoemack, P., Ren, X., Storey, M., Jefferies, S., Smit, E., Hadfield, J., Kenny, A., Jelley, L., Sporle, A., McNeill, A., Reynolds, G. E., Mouldey, K., Lowe, L., Sonder, G., Drummond, A. J., Huang, S., Welch, D., Holmes, E. C., French, N., Simpson, C. R., & de Ligt, J. (2021). Genomic Evidence of In-Flight Transmission of SARS-CoV-2 Despite Predeparture Testing. Emerg Infect Dis, 27(3), 687–693. https://doi.org/10.3201/eid2703.204714

WHO. (2009). Case management of Influenza A(H1N1) in air transport. https://www.who.int/csr/resources/publications/swineflu/air_transport/en/

WHO. (2020). Operational planning guidance to support country preparedness and response. World Health Organization. Retrieved 3/23/2021 from https://www.who.int/publications/i/item/draft-operational-planning-guidance-for-un-country-teams

Yang, N., Shen, Y., Shi, C., Ma, A. H. Y., Zhang, X., Jian, X., Wang, L., Shi, J., Wu, C., Li, G., Fu, Y., Wang, K., Lu, M., & Qian, G. (2020). In-flight transmission cluster of COVID-19: a retrospective case series. Infect Dis (Lond), 52(12), 891–901. https://doi.org/10.1080/23744235.2020.1800814

Zhang, J., Li, J., Wang, T., Tian, S., Lou, J., Kang, X., Lian, H., Niu, S., Zhang, W., Jiang, B., & Chen, Y. (2020). Transmission of SARS-CoV-2 on Aircraft In (Pre-print ed.): SSRN.

Zhang, X. A., Fan, H., Qi, R. Z., Zheng, W., Zheng, K., Gong, J. H., Fang, L. Q., & Liu, W. (2020). Importing coronavirus disease 2019 (COVID-19) into China after international air travel. Travel Med Infect Dis, 35, 101620. https://doi.org/10.1016/j.tmaid.2020.101620

