## Appendix 1 for "Transmission of SARS-CoV-2 associated with aircraft travel: a systematic review (Version 1)"

Protocol for a living evidence review

Rosca EC, ^1^ Heneghan C, ^2^ Spencer EA, ^2^ Brassey J, ^3^ Plüddemann A, ^2^ Onakpoya IJ, ^2^ Evans D, ^4^ Conly JM, ^5^ Jefferson T. ^2^

Affiliations

1. Victor Babes University of Medicine and Pharmacy of Timisoara, Romania
2. University of Oxford, UK
3. Trip Database Ltd, UK
4. Li Ka Shing Institute of Virology and Dept. of Medical Microbiology & Immunology, University of Alberta
5. University of Calgary and Alberta Health Services, Calgary, Canada.

**Keywords**

COVID-19; SARS-CoV-2; Transmission; Aircraft.

**Background:**

COVID‑19 is a new disease, distinct from other diseases caused by coronaviruses, such as Severe Acute Respiratory Syndrome (SARS) and Middle East Respiratory Syndrome (MERS). The SARS-CoV-2 virus spreads rapidly, and the modes of transmission are not completely understood .

National and international authorities, including WHO, have been working urgently to coordinate the response with the swift development of prevention, control, and management measures on many fronts. Their overarching aim is to control the current pandemics by suppressing transmission of SARS-CoV-2 and preventing associated morbidity and mortality [WHO 2020]. Nonetheless, the transmission of the virus and the many aspects of the disease are not completely understood. Therefore, the public health measures for restricting transmission are based on the best available information [EASA 2020].

Aircraft travel was reported to be associated with the spread of pathogen agents through infected passengers, possibly through in-flight transmission. The increased likelihood of transmission of infectious diseases could be due to the high number of passengers, often in close proximity to each other. Therefore, the pathogens may spread through multiple routes of transmission. As in other enclosed or semi-closed settings, the in-flight transmission of viruses is promoted by close contact or contact with contaminated surfaces [Leitmeyer 2016; Leitmeyer 2011; ECDC 2014]. The transmission risk depends on proximity to an index case, and contact among passengers, movement of the passengers or crew, and fomites [ECDC 2017].

Currently, there are several disease-specific guidance recommendations for case management in air transport for various pathogens, elaborated by international authorities like the WHO and the European Centre for Disease Prevention and Control [WHO 2009; ECDC 2014].

The SARS-CoV-2 is a novel virus. Therefore, the initial transmission models of spread associated with air travel are based on what is known on the dynamics of other respiratory viral infections, mainly coronaviruses, and influenza. However, an important aspect of the current uncertainties consists of the modes and circumstances of transmission of newly identified agents. Therefore, research is ongoing to understand SARS-CoV-2 modes of transmission, complemented with a growing number of new publications. Consequently, there is a need to continuously and systematically perform reviews of studies with the latest data available in order to inform recommendations using the most up-to-date information.

**Objectives**

We aim to provide a rapid summary and evaluation of relevant data on the transmission of SARS-CoV-2 associated with aircraft travel, report important policy implications, and highlight research areas that are urgently needed. This transmission area includes airborne, contact and droplet, fomite, and orofecal.

**Modes of transmission for SARS-CoV-2** (WHO 2020, WHO 2014)

The virus can spread from an infected person’s mouth or nose in small liquid particles when the person coughs, sneezes, sings, breathes heavily, or talks. The liquid particles are of different sizes (ranging from larger ‘respiratory droplets’ to smaller ‘aerosols.’).

● Respiratory droplets are >5-10 μm in diameter. Respiratory droplets that include viruses can reach the mouth, nose, or eyes of a susceptible person and can result in infection.

**●**	Respiratory droplets <5μm in diameter are referred to as droplet nuclei or aerosols. Airborne transmission is the spread of an infectious agent caused by disseminating aerosols that remain infectious when suspended in the air over long distances and time.

**●** Close contact transmission (typically within 1 meter) occurs with inhalation of SARS-CoV-2 or inoculation with the virus through the mouth, nose, or eyes**.**

**●**	Transmission through fomites (surfaces and objects that may be contaminated with a viable virus, in the immediate environment around the infected person) **.**

**●**	Orofecal transmission occurs where the virus in fecal particles can pass from one person to the mouth of another. The main causes include lack of adequate sanitation and poor hygiene practices. Another form of orofecal transmission consists of fecal contamination of food.

**Subgroups:**

If feasible, the studies on the in-flight transmission of SARS-CoV-2 will form a subgroup of a broader systematic review on transmission dynamics of COVID-19 [Jefferson 2020]. We will report the evidence from studies with the results of reverse transcriptase-polymerase chain reaction (RT-PCR) where reported by cycle threshold (Ct), time from symptom onset and live culture of SARS-CoV-2. Evidence from studies comparing culture with other means of diagnosis is not usually mode of transmission-specific. Updates of the culture review will be carried out alongside and in parallel with the mode of transmission study extraction for all modes.

**Methods**

This protocol is developed based on a previous protocol for a series of systematic reviews on the evidence on transmission dynamics of COVID-19 [Jefferson 2020].

**Search Strategy**

We will search the following electronic databases: LitCovid, medRxiv, Google Scholar, and the WHO Covid-19 database. Searches will be updated approximately every month; if the volume of new studies will be significant, searching will be performed every fortnight. Search terms are (coronavirus OR covid-19 OR SARS-CoV-2( AND (aircraft OR airplane OR flight) AND transmission. Because WHO Covid-19 Database and LitCovid are specific Covid-19 databases, there will be no requirement to add a search string to identify Covid-19 articles. In addition, reference lists of relevant articles, including reviews, will be screened for additional relevant studies.

**Study inclusion and exclusion**

We will include studies reporting onboard transmission of COVID-19 from passengers and crew to passengers or crew. We will consider any potential transmission mode, including droplet, airborne, fomite, fecal-oral, or other. Studies can be observational, including case series, ecological, or prospective; or interventional, including randomised trials and clinical reports, outbreak reports, case-control studies, experimental studies, non-predictive modelling.

We will include studies on factors influencing transmission, such as infectivity of the index case (pre-symptomatic, asymptomatic or symptomatic, with or without mask), the susceptibility of passengers (previous COVID-19 infection or vaccination, wearing or not masks), and effectiveness of exposure (proximity to the index case, duration of exposure, technical specifications of the airplane, quality of cabin air). Studies incorporating models to describe observed data are included. Studies reporting solely predictive modelling are excluded.

Pre-symptomatic transmission is defined as transmission from people who are infected and shedding virus but have not yet developed symptoms [WHO 2020].

Asymptomatic transmission is defined as transmission from people infected with SARS-CoV-2 who never develop symptoms [WHO 2020].

**Data extraction**

We will include publication details (authors, year, country); study type; flight characteristics (origin and destination of the aircraft, flight duration, technical specifications of the airplane, ground delays, and information on ventilation system); data on the index cases (number, age, gender, country of residence or nationality, seating, if they wore or not masks, symptoms during the flight, laboratory confirmation of diagnosis); details on contact tracing (definition of contact, secondary cases demographic data, symptoms, laboratory confirmation, contact tracing strategy, methods used to identify contacts, methods used for contacting contacts, total number of contacts identified, the total number of successfully traced contacts, the seating of contacts in relation to the index case, immunological status and if they wore or not masks); exposure of primary and secondary cases (before, during, and after the flight); conclusion on disease transmission (the number of cases/number of contacted passengers, and crew excluding index cases), interventions used, and funding source for the study.

Study data will be extracted into data extraction templates Table 1 and Appendix 1 (study characteristics), Table 2 (methodological quality of studies), and a table with a summary of main findings. References will be included in alphabetical order as a web appendix that facilitates updating. We will follow PRISMA reporting guidelines.

Data extraction will be performed by one author and checked by a second author. In case of disagreement, a third author will arbitrate.

**Quality assessment**

We will assess the quality of included studies based on a modified QUADAS-2 tool using five criteria. For the in-flight transmission studies, we will investigate the following aspects:

(1) a clearly defined setting, with data on the aircraft type, location of index cases, and secondary cases (2) demographic characteristics (including age and gender) and sampling procedures adequately described, with data on the day of the sampling procedure and data on symptoms (including onset day) (3) follow-up strategy and duration sufficient for the outcomes; we will consider as adequate a comprehensive follow-up strategy, for at least 14 days

(4) the transmission outcomes assessed adequately; the studies should report the following data on secondary cases: demographic data, clinical data (with the day of onset of symptoms), and paraclinical data (RT-PCR with Ct <25; GS; viral cultures) with the day of the procedure

(5) main biases that are threats to validity taken into consideration, follow-up of >80% of passengers and crew, and exclusion of alternative exposures

Quality assessment will be performed by one author and checked by a second author. If a disagreement is not solved by discussion, a third author will arbitrate.

For studies that generate the hypothesis of onboard COVID-19 transmission, we will assess the strength of evidence for each study depending on the methods used to investigate the SARS-CoV-2 transmission [Jefferson 2021].

For the non-human studies, we will investigate the following aspects:

(1) Is the description of methods with sufficient detail to replicate
(2) Is the sample source clearly described
(3) Are the analyses and reporting appropriate
(4) Is the bias dealt with
(5) Are there any applicability concerns?

**Data synthesis and reporting**

The outcome consists of the onboard transmission of COVID-19. We will provide a narrative summary of the review and report the outcomes, including quantitative estimates where feasible and relevant. We will report the detection of a live culture of SARS-CoV-2 when reported (see also subgroups). Where possible, compatible datasets may be pooled for meta-analysis. We may write to authors for clarification of data and also report research and policy implications.

**Continual data release**

Summary descriptions of important, relevant research papers identified are summarised in the tracker and corresponding folders in an ongoing manner. In addition, as important new data accumulates, we produce a report as an individual rapid review and aim to make all our work available by depositing the review findings on the Oxford Research Archive.

**References:**

European Centre for Disease Prevention and Control (ECDC). Risk assessment guidelines for infectious diseases transmitted on aircraft (RAGIDA) - Influenza. Available at <https://www.ecdc.europa.eu/en/publications-data/risk-assessment-guidelines-infectious-diseases-transmitted-aircraft-ragida>

Jefferson T, Plüddemann A, Spencer EA, Brassey J, Heneghan C. The evidence on transmission dynamics of COVID-19: protocol for a series of Systematic Reviews. Available at <https://www.cebm.net/evidence-synthesis/transmission-dynamics-of-covid-19/>

Jefferson, T.; Heneghan, C.; Spencer, E.; Brassey, J.; Pluddeman, A.; Onakpoya, I.; Evans, D.; Conly, J. A Hierarchical Framework for Assessing Transmission Causality of Respiratory Viruses . *Preprints* **2021**, 2021040633 (doi: 10.20944/preprints202104.0633.v1).

Leitmeyer K. European risk assessment guidance for infectious diseases transmitted on aircraft–the RAGIDA project. Euro Surveill. 2011;16(16):19845.

Leitmeyer K, Adlhoch C. Review Article: Influenza Transmission on Aircraft: A Systematic Literature Review. Epidemiology. 2016; 27(5):743-51. doi: 10.1097/EDE.0000000000000438.

World Health Organization. WHO technical advice for case management of Influenza A(H1N1) in air transport. May 13, 2009; Available at: <https://www.who.int/csr/resources/publications/swineflu/air_transport/en/>

World Health Organization. Mask use in the context of COVID-19: interim guidance, 1 December 2020: [Mask use in the context of COVID-19: interim guidance, 1 December 2020 (who.int)](https://apps.who.int/iris/handle/10665/337199)

World Health Organization. Infection Prevention and Control of Epidemic-and Pandemic-prone Acute Respiratory Infections in Health Care. Geneva; 2014 (available at <https://apps.who.int/iris/bitstream/handle/10665/112656/9789241507134_eng.pdf;jsessionid=41AA684FB64571CE8D8A453C4F2B2096?sequence=1>).

Public Health England. Understanding cycle threshold (Ct) in SARS-CoV-2 RT-PCR. A guide for health protection teams. 2020. [https://www.gov.uk/government/publications/cycle-threshold-ct-in-sars-cov-2-rt-pc](https://www.gov.uk/government/publications/cycle-threshold-ct-in-sars-cov-2-rt-pcr)r

World Health Organization. Operational planning guidance to support country preparedness and response. Geneva: World Health Organization; 2020 <https://www.who.int/publications/i/item/draft-operational-planning-guidance-for-un-country-teams>

World Health Organization. Information Notice for IVD Users 2020/05. Nucleic acid testing (NAT) technologies that use polymerase chain reaction (PCR) for detection of SARS-CoV-2. <https://www.who.int/news/item/20-01-2021-who-information-notice-for-ivd-users-2020-05>
