## Appendix 4 for "Transmission of SARS-CoV-2 associated with aircraft travel: a systematic review (Version 1)"

**List of Excluded studies**

Bielecki M, Patel D, Hinkelbein J, Komorowski M, Kester J, Ebrahim S, Rodriguez-Morales AJ, Memish ZA, Schlagenhauf P. Reprint of: Air travel and COVID-19 prevention in the pandemic and peri-pandemic period: A narrative review. Travel Med Infect Dis. 2020;38:101939. doi: 10.1016/j.tmaid.2020.101939. – Narrative Review

Freedman DO, Wilder-Smith A. In-flight transmission of SARS-CoV-2: a review of the attack rates and available data on the efficacy of face masks. J Travel Med. 2020;27(8):taaa178. doi: 10.1093/jtm/taaa178. – Narrative Review

Hu M, Wang J, Lin H, Ruktanonchai CW, Xu C, Meng B, et al. Transmission risk of SARS-CoV-2 on airplanes and high-speed trains. medRxiv. 2020:2020.12.21.20248383. doi: <https://doi.org/10.1101/2020.12.21.20248383> - Modeling

Luo G, McHenry ML, Letterio JJ. Estimating the prevalence and risk of COVID-19 among international travelers and evacuees of Wuhan through modeling and case reports. PLoS One. 2020;15(6):e0234955. doi: 10.1371/journal.pone.0234955. – Modeling

Swadi, Tara; L. Geoghegan, Jemma; Devine, Tom; McElnay, Caroline; Shoemack, Phil; Ren, Xiaoyun; et al. (2020): A case study of extended in-flight transmission of SARS-CoV-2 en route to Aotearoa New Zealand. Institute of Environmental Science and Research. Preprint. <https://doi.org/10.26091/ESRNZ.13257914.v1>
