## Appendix 5 for "Transmission of SARS-CoV-2 associated with aircraft travel: a systematic review (Version 1)"

**Web table. References: Aircraft transmission of SARs-CoV-2**

References Web supplement search up to 27 /01/2021 (n= 20 primary studies; 0 systematic review):

**References: Included Studies (n=20)**

*Case reports, case series, cohort studies*

Bae SH, Shin H, Koo HY, Lee SW, Yang JM, Yon DK. Asymptomatic Transmission of SARS-CoV-2 on Evacuation Flight. Emerg Infect Dis. 2020;26 (11):2705-2708. doi: 10.3201/eid2611.203353.

Chen J, He H, Cheng W, Liu Y, Sun Z, Chai C, Kong Q, Sun W, Zhang J, Guo S, Shi X, Wang J, Chen E, Chen Z. Potential transmission of SARS-CoV-2 on a flight from Singapore to Hangzhou, China: An epidemiological investigation. Travel Med Infect Dis. 2020; 36:101816. doi: 10.1016/j.tmaid.2020.101816.

Choi EM, Chu DKW, Cheng PKC, Tsang DNC, Peiris M, Bausch DG, Poon LLM, Watson-Jones D. In-Flight Transmission of SARS-CoV-2. Emerg Infect Dis. 2020; 26(11):2713-2716. doi: 10.3201/eid2611.203254.

Eldin C, Lagier JC, Mailhe M, Gautret P. Probable aircraft transmission of Covid-19 in-flight from the Central African Republic to France. Travel Med Infect Dis. 2020; 35:101643. doi: 10.1016/j.tmaid.2020.101643

Hoehl S, Karaca O, Kohmer N, Westhaus S, Graf J, Goetsch U, Ciesek S. Assessment of SARS-CoV-2 Transmission on an International Flight and Among a Tourist Group. JAMA Netw Open. 2020; 3(8): e2018044. doi: 10.1001/jamanetworkopen.2020.18044.

Khanh NC, Thai PQ, Quach HL, Thi NH, Dinh PC, Duong TN, Mai LTQ, Nghia ND, Tu TA, Quang N, Quang TD, Nguyen TT, Vogt F, Anh DD. Transmission of SARS-CoV 2 During Long-Haul Flight. Emerg Infect Dis. 2020; 26(11):2617-2624. doi: 10.3201/eid2611.203299.

Kong D, Wang Y, Lu L, Wu H, Ye C, Wagner AL, Yang J, Zheng Y, Gong X, Zhu Y, Jin B, Xiao W, Mao S, Jiang C, Lin S, Han R, Yu X, Cui P, Fang Q, Lu Y, Pan H. Clusters of 2019 coronavirus disease (COVID-19) cases in Chinese tour groups. Transbound Emerg Dis. 2020: 10.1111/tbed.13729. doi: 10.1111/tbed.13729.

Murphy N, Boland M, Bambury N, Fitzgerald M, Comerford L, Dever N, O'Sullivan MB, Petty-Saphon N, Kiernan R, Jensen M, O'Connor L. A large national outbreak of COVID-19 linked to air travel, Ireland, summer 2020. Euro Surveill. 2020; 25(42):2001624. doi: 10.2807/1560-7917.ES.2020.25.42.2001624.

Ng OT, Marimuthu K, Chia PY, Koh V, Chiew CJ, De Wang L, Young BE, Chan M, Vasoo S, Ling LM, Lye DC, Kam KQ, Thoon KC, Kurupatham L, Said Z, Goh E, Low C, Lim SK, Raj P, Oh O, Koh VTJ, Poh C, Mak TM, Cui L, Cook AR, Lin RTP, Leo YS, Lee VJM. SARS-CoV-2 Infection among Travelers Returning from Wuhan, China. N Engl J Med. 2020; 382(15):1476-1478. doi: 10.1056/NEJMc2003100.

Nir-Paz R, Grotto I, Strolov I, Salmon A, Mandelboim M, Mendelson E, Regev-Yochay G. Absence of in-flight transmission of SARS-CoV-2 likely due to use of face masks on board. J Travel Med. 2020; 27(8): taaa117. doi: 10.1093/jtm/taaa117.

Park JH, Jang JH, Lee K, Yoo SJ, Shin H. COVID-19 Outbreak and Presymptomatic Transmission in Pilgrim Travelers Who Returned to Korea from Israel. J Korean Med Sci. 2020; 35(48):e424. doi: 10.3346/jkms.2020.35.e424.

Pavli A, Smeti P, Hadjianastasiou S, Theodoridou K, Spilioti A, Papadima K, Andreopoulou A, Gkolfinopoulou K, Sapounas S, Spanakis N, Tsakris A, Maltezou HC. In-flight transmission of COVID-19 on flights to Greece: An epidemiological analysis. Travel Med Infect Dis. 2020; 38:101882. doi: 10.1016/j.tmaid.2020.101882.

Schwartz KL, Murti M, Finkelstein M, Leis JA, Fitzgerald-Husek A, Bourns L, Meghani H, Saunders A, Allen V, Yaffe B. Lack of COVID-19 transmission on an international flight. CMAJ. 2020;192(15): E410. doi: 10.1503/cmaj.75015.

Speake H, Phillips A, Chong T, Sikazwe C, Levy A, Lang J, Scalley B, Speers DJ, Smith DW, Effler P, McEvoy SP. Flight-Associated Transmission of Severe Acute Respiratory Syndrome Coronavirus 2 Corroborated by Whole-Genome Sequencing. Emerg Infect Dis. 2020; 26(12):2872-2880. doi: 10.3201/eid2612.203910.

Swadi T, Geoghegan JL, Devine T, McElnay C, Sherwood J, Shoemack P, Ren X, Storey M, Jefferies S, Smit E, Hadfield J, Kenny A, Jelley L, Sporle A, McNeill A, Reynolds GE, Mouldey K, Lowe L, Sonder G, Drummond AJ, Huang S, Welch D, Holmes EC, French N, Simpson CR, de Ligt J. Genomic Evidence of In-Flight Transmission of SARS-CoV-2 Despite Predeparture Testing. Emerg Infect Dis. 2021; 27(3):687-693. doi: 10.3201/eid2703.204714.

Yang N, Shen Y, Shi C, Ma AHY, Zhang X, Jian X, Wang L, Shi J, Wu C, Li G, Fu Y, Wang K, Lu M, Qian G. In-flight transmission cluster of COVID-19: a retrospective case series. Infect Dis (Lond). 2020; 52(12):891-901. doi: 10.1080/23744235.2020.1800814.

Zhang, Jinjun and Li, Jianren and Wang, Tianbing and Tian, Sijia and Lou, Jing and Kang, Xuqin and Lian, Huixin and Niu, Shengmei and Zhang, Wenzhong and Jiang, Baoguo and Chen, Yuguo, Transmission of SARS-CoV-2 on Aircraft (4/23/2020). Available at SSRN: https://ssrn.com/abstract=3586695 or<http://dx.doi.org/10.2139/ssrn.3586695>.

Zhang XA, Fan H, Qi RZ, et al. Importing coronavirus disease 2019 (COVID-19) into China after international air travel. Travel Med Infect Dis. 2020; 35:101620. doi: 10.1016/j.tmaid.2020.101620.

*Wastewater studies*

Ahmed W, Bertsch PM, Angel N, Bibby K, Bivins A, Dierens L, Edson J, Ehret J, Gyawali P, Hamilton KA, Hosegood I, Hugenholtz P, Jiang G, Kitajima M, Sichani HT, Shi J, Shimko KM, Simpson SL, Smith WJM, Symonds EM, Thomas KV, Verhagen R, Zaugg J, Mueller JF. Detection of SARS-CoV-2 RNA in commercial passenger aircraft and cruise ship wastewater: a surveillance tool for assessing the presence of COVID-19 infected travellers. J Travel Med. 2020; 27(5): taaa116. doi: 10.1093/jtm/taaa116.

Albastaki A, Naji M, Lootah R, Almeheiri R, Almulla H, Almarri I, Alreyami A, Aden A, Alghafri R. First confirmed detection of SARS-COV-2 in untreated municipal and aircraft wastewater in Dubai, UAE: The use of wastewater based epidemiology as an early warning tool to monitor the prevalence of COVID-19. Sci Total Environ. 2021;7 60:143350. doi: 10.1016/j.scitotenv.2020.143350.
