## Appendix 6 for "Transmission of SARS-CoV-2 associated with aircraft travel: a systematic review (Version 1)"

**Appendix 2.** Comparison of the studies that very likely report on the same flight. The similarities are highlighted in red.

| Data | Chen 2020 | Yang 2020 |
| --- | --- | --- |
| Route | Singapore to Hangzhou | Singapore to Hangzhou |
| Flight date | 24 January 2020 | 24 January 2020 |
| Flight duration | 4 hours 50 min – arrival at 9:40 pm | 5 hours – arrival at 10:00 pm |
| Aircraft type | Boeing 787-9 | N/R |
| Index case definition | Passenger who had a SARSCoV- 2 infection confirmed by RT-PCR testing of a throat swab sample, and had symptoms of typical upper respiratory tract infection; Asymptomatic cases - positive RT-PCR test results but without any COVID-19 symptoms. The date of the positive RT-PCR test result was used as the onset date | Px with symptoms during flight, with positive RT-PCR. |
| Contact definition | All passengers on the flight | Px on the flight. The incubation period was defined as the time from the flight arrival to the onset of illness. |
| Number of px | 335 px, 11 crew | 325 px and crew |
| Covid 19 cases | 26 F – symptomatic one day before flight | 26F – symptomatic D0 |
|  | 45 M – symptomatic D0 – among index cases | 45M - symptomatic D0 – index case |
|  | 29F – symptomatic D2 | 20F – symptomatic D3 |
|  | 52F – symptomatic D3 | 52F – symptomatic D3 |
|  | 45F – symptomatic D2 | 45F – symptomatic D2 |
|  | 33F – symptomatic one day before flight | 33F – symptomatic D1 |
|  | 36F – symptomatic D3 | - |
|  | 33F – symptomatic D7 | 33F – symptomatic D8 |
|  | 32M – symptomatic D9 | 32M -symptomatic D8 |
|  | 44M – symptomatic D9 | - |
|  | 44F – asymptomatic – RT-PCR positive on D2 | - |
|  | 44M – asymptomatic - RT-PCR positive on D2 | - |
|  | 26F - asymptomatic - RT-PCR positive on D2 | 25F – asymptomatic, RT-PCR positive on D15 |
|  | 28M – asymptomatic - RT-PCR positive on D2 | - |
|  | 27F – asymptomatic - RT-PCR positive on D2 | 27F – asymptomatic, RT-PCR positive on D15 |
|  | Total 10 symptomatic, 6 asymptomatic | Total 8 symptomatic, 2 asymptomatic |
| Case classification | 15 index cases, 1 secondary case | 1 index case, 9 secondary cases |
| Follow-up | All the px on the flight. Px with symptoms were sent to the hospital  Quarantine of all px, 14 days;  Interview by telephone or face to face | 10 px (that were admitted to the hospital); Authors state that in addition, another 2 px tested positive, but they failed to interview them (Could be 36F and 44M from the study of Yang).  Retrospective study, based on medical records |
| Intervention | Px with symptoms were sent to hospital (n=10) | Hospitalization |

We also found some additional data on a flight from Singapore to Hangzhou on 24 January 2020 in the news magazines at that time:

<https://fortune.com/2020/07/28/unmasked-airline-passenger-rules-masks/>

[All passengers & crew on Scoot flight from S'pore to China isolated, 1 sent for further blood test - Mothership.SG - News from Singapore, Asia and around the world](https://mothership.sg/2020/01/scoot-wuhan-singapore/)

In the article from the Mothership website, we found the Scoot declaration: "Scoot has always operated Hangzhou flights with widebody Boeing 787 aircraft, and TR188 on 24 January 2020 was carrying 314 passengers in total."

However, in the same article, it is stated that " According to a Weibo post by *CCTV News*, a Scoot flight TR188 from Singapore landed in Hangzhou at around 10pm on Jan. 24. The flight allegedly had 335 passengers, including 116 Wuhan natives".

The article of Chen 2020 mentions 335 passengers. On the other hand, the report of Yang 2020 states there were 325 passengers.

Furthermore, we searched flight websites. On the Flightera website we found the actual flight, with some details:

<https://www.flightera.net/en/flight_details/Scoot-Singapore-Hangzhou/TR188/WSSS/2020-01-24>

Therefore, we have found data suggesting both articles report the same flight, namely a specific repatriation flight of Wuhan residents from Singapore. In our opinion, Yang et al. reported the investigation of the patients admitted to their hospital, whereas Chen et al. performed a more thorough investigation, with much detailed information.

The present papers indicate duplicate publications unknown to each other. Furthermore, they are an example of how the same data can be differently interpreted, one e suggesting major in-flight transmission and the other the opposite.
